# Modelling serological cross-reactivity to disentangle the dynamics of West Nile and Usutu viruses in an emerging area

**DOI:** 10.64898/2026.04.07.26350295

**Authors:** Jonathan Bastard, Camille Migné, Teheipuaura Helle, Estelle Agneray, Clément Bigeard, Yasmine Boudjadi, Manon Chevrier, Marine Dumarest, Mathilde Gondard, Sandra Martin-Latil, Laure Mathews-Martin, Thierry Petit, Thomas Charpentier, Hanae Pouillevet, Benoit Durand, Raphaëlle Métras, Gaëlle Gonzalez

## Abstract

Zoos may serve as sentinel sites for zoonotic vector-borne diseases. West Nile virus (WNV) and Usutu virus (USUV) are closely related orthoflaviviruses transmitted between *Culex* mosquitoes and a bird reservoir. Both viruses can also infect mammals, including humans, where they may cause symptoms and, more rarely, hospitalization and death. However, serological cross-reactivity between WNV and USUV complicates their differential diagnosis. Here, we aimed to reconstruct the dynamics of emergence of WNV in a zoo located in a newly affected area in Europe, using ELISA and Virus Neutralization Test (VNT) serological analysis of 1707 animal sera collected between 2015 and 2024. Combining this data in a model accounting for cross-reactivity with USUV, we estimated yearly forces of infection (FOI) by both viruses, and thus found that WNV likely circulated in the area one year prior to the first cases reported to the passive surveillance system. Our results also showed that, in the zoo, mammals and reptiles had a lower risk of infection than birds (relative risk of 0.14 [0.05; 0.28]), and that the exposure of birds to water (aquatic lifestyle or proximity to stagnant water) affected the risk. Finally, we estimated diagnosis parameters, including the sensitivity of the VNT (80.4% [76.5%; 84.3%]), the expected VNT titer value, and the level of serological cross-reactivity between viruses during the VNT. To conclude, our modelling framework allowed to disentangle the co-circulation of two closely related viruses, a crucial point in ensuring the reliable sentinel surveillance of these vector-borne zoonotic pathogens.

## Introduction

Zoos have been advocated as possible sentinel sites for monitoring the circulation of zoonotic infectious diseases for several reasons (Van Leeuwen et al. 2023; Hernandez-Colina et al. 2024). First, zoos provide a controlled environment and easy sampling of animals, for instance as part of routine veterinary health operations. Second, zoo individuals generally have well-documented records of birth, death, enclosure locations and movements between institutions (Che-Castaldo et al. 2019). Third, such facilities often host a large diversity of taxa in close proximity, including species with various susceptibility to infection or various attractiveness to vectors (Van Leeuwen et al. 2023; Roche et al. 2015). Fourth, zoos are frequently close to human dwellings, which is an asset for sentinel sites (Neo and Tan 2017).

West Nile virus (WNV) and Usutu virus (USUV) are widely spread orthoflaviviruses, belonging to the Japanese encephalitis virus antigenic complex, and are both transmitted between *Culex* mosquitoes and birds (Simonin 2024). Besides this vector-related transmission, previous studies suggested the existence of a non-vector, possibly orofecal, transmission route between birds for WNV (Banet-Noach et al. 2003; Hinton et al. 2015; Hartemink et al. 2007) and USUV (Kuchinsky et al. 2021; Benzarti et al. 2020). Both viruses can also infect mammals and, while most often asymptomatic, can cause neurological symptoms in humans, which are considered dead-end hosts (Angeloni et al. 2023; Simonin 2024). Since WNV seems to be more virulent than USUV in humans (Marshall et al. 2025), it is important to differentiate their areas of circulation. Both viruses originate from Africa and may spread through migratory corridors. In Europe, WNV was historically mostly detected in Central and Eastern Europe, and around the Mediterranean Basin (Italy, South of France, Greece, among others). However, its geographical range has been expanding in Europe, with detections in Germany in 2018, in the Netherlands in 2020, and Western and Northern regions of France starting from 2022 (Chevalier et al. 2025; ECDC 2025). USUV was found to circulate in Italy as early as 1996. Since then, it has been reported to cause bird mortalities and sporadic human clinical cases in several European countries (Angeloni et al. 2023; Simonin 2024). In France, USUV was first described in birds in 2015, in the eastern part of the country (Lecollinet et al. 2016). The intensity of circulation of both viruses was shown to present year-to-year variations (Chevalier et al. 2025; Angeloni et al. 2023; ECDC 2025).

The Nouvelle-Aquitaine region, on the Atlantic coast of France near Bordeaux, can be considered as a WNV emergence area. Indeed, while this virus was historically absent from the region, three equine clinical cases were first notified in 2022 (Chevalier et al. 2025). However, WNV infections may have occurred in the area before this first detection. In 2023, the affected geographical area within the region expanded and multiple clinical cases were notified in humans, horses and birds. In particular, WNV was shown to cause mortality in birds, mostly Chilean (*Phoenicopterus chilensis*) and Cuban (*Phoenicopterus ruber*) flamingoes, kept in a local zoo (Duvignaud et al. 2024), where a serum bank had been previously implemented. This provided an opportunity for retrospectively investigating biomarkers of this arbovirus’ circulation in zoo animals’ sera.

Serosurveys for WNV and/or USUV have been previously led in zoos across the world. They allowed to document viral emergence in new geographical areas, such as WNV in North America starting from 1999 (Ludwig et al. 2002; Farfán-Ale et al. 2006). They also provided seroprevalence estimates for these pathogens, showing that they could infect many species (Ludwig et al. 2002; Farfán-Ale et al. 2006; Cano-Terriza et al. 2015; Caballero-Gómez et al. 2020; Constant et al. 2020; Kvapil et al. 2021). However, serosurveys’ results and their interpretations in terms of diagnosis and surveillance may be jeopardized by the occurrence of serological cross-reactivity in arboviruses, especially between orthoflaviviruses (Madere et al. 2025; Čabanová et al. 2023; de Bellegarde de Saint Lary et al. 2023; Lustig et al. 2018), highlighting the need for adequate statistical methods to address this issue (Hozé et al. 2020, 2021; O’Driscoll et al. 2024).

Here, we had the objective to reconstruct the emergence of WNV in a zoo located in a newly affected geographical area in Europe by analyzing (i) serological and demographic (exposure) data available from multiple zoo species during 10 years, using (ii) a modelling framework that allowed to account for the serological cross-reactivity between WNV and USUV. We found that WNV likely circulated undetected in the area, one year prior to being reported by the passive surveillance systems in humans and animals, and discuss implications for future arboviral emergences.

## Material and Methods

### Sera and associated data

Serological data was obtained thanks to a serum bank implemented in La Palmyre zoo (Les Mathes, Nouvelle-Aquitaine, France). Animal sera were sampled by convenience when they were handled for routine veterinary operations or vaccination. In this study, we analyzed all sera collected in the zoo between January 2015 and November 2024, i.e. 1707 sera from 940 animals of 84 species, including 18 bird species, 60 mammal species and 6 reptile species (Figure 1 and Supplementary Table S1). A median of 2 samples was collected per individual (interquartile range: [1; 2]).

**Figure 1.**
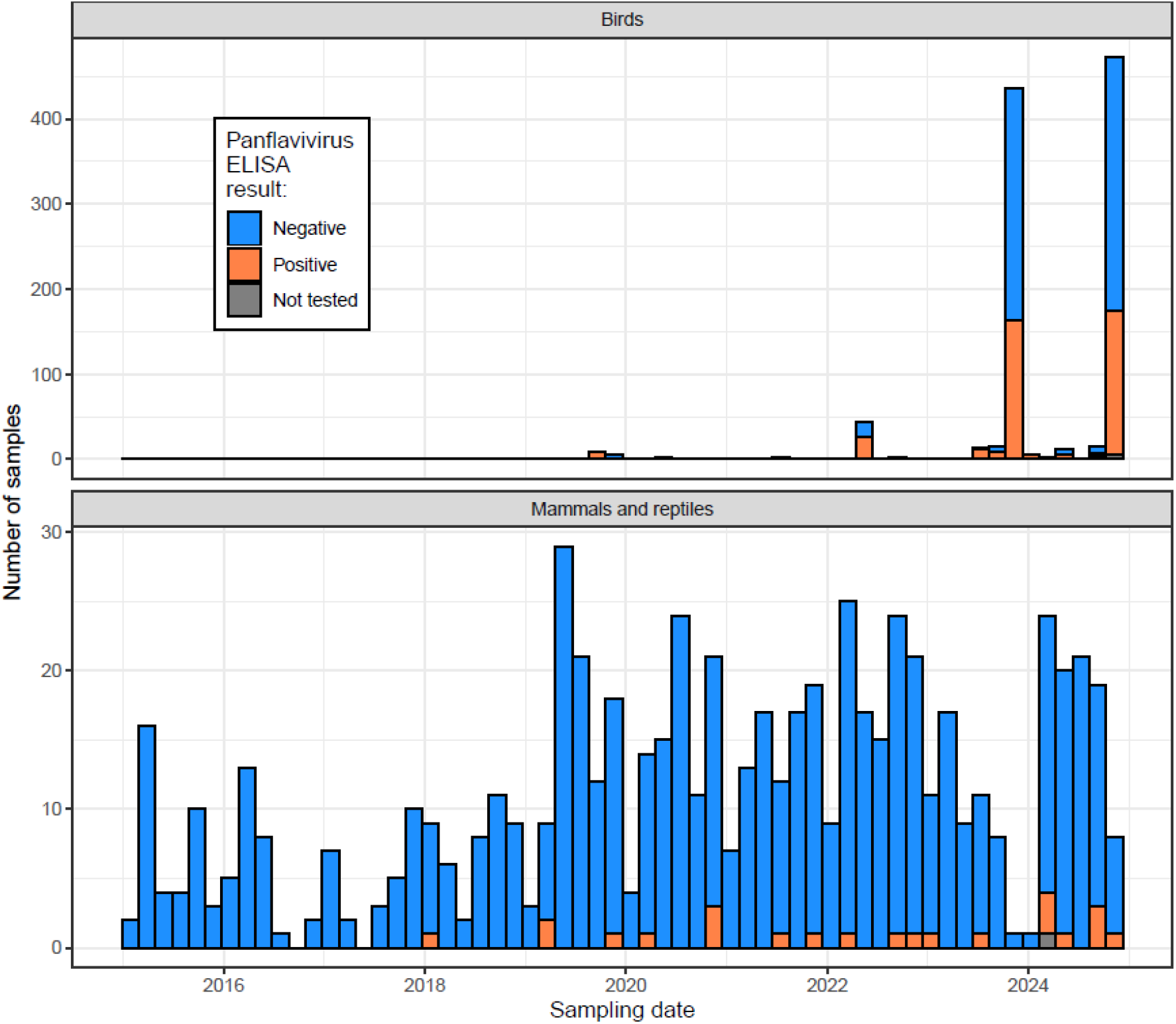
Number of serum samples collected in La Palmyre zoo (Les Mathes, Nouvelle-Aquitaine, France) between January 2015 and November 2024 (1707 sera from 940 animals), in birds (upper panel), mammals and reptiles (lower panel), and their Panflavivirus ELISA test results (positive or negative). All samples but 9 were analyzed using a Panflavivirus ELISA test. For model analysis, ELISA doubtful results were considered as positive.

On average between 2016 and 2024, the total number of animals kept at the zoo was 1277, including 691 birds of 27 species, 501 mammals of 68 species, and 85 reptiles of 15 species (see the detail by species in Supplementary Table S2).

For each individual, information was recorded on its date of birth or arrival at the zoo and, when applicable, its date of death or departure from the zoo. We were therefore able to retrieve the number of days each individual was present at the zoo prior to serological sampling. We only included in the exposure time the periods between May 1^st^ and November 30^th^ of each year (the mosquito season). Moreover, we assessed the level of birds’ exposure to waterbodies within the zoo based on (i) the presence of stagnant water in their enclosure in summer nights – because *C. pipiens* vectors tend to bite at night (Veronesi et al. 2012) – and (ii) the importance of water in the species’ feeding behavior or the role of frequent water immersion or bathing in its ecology. Overall, we classified individuals into the following categories: “Mammals and reptiles” (380/940 individuals), “Birds with no water exposure” (16/940 individuals), “Birds with aquatic lifestyle only” (127/940 individuals), “Birds with proximity to stagnant water in summer nights only” (14/940 individuals) and “Birds with both water exposures” (403/940 individuals) (Supplementary Table S1).

### Serological screening

All samples but 9 were tested for the presence of anti-orthoflavivirus IgG antibodies with a competition Panflavivirus Enzyme-Linked Immuno-Sorbent Assay (ELISA) test. We used the kit ID Screen® Flavivirus Competition (Innovative Diagnostic, Montpellier, France) according to supplier’s instructions. The results are expressed in %S/N. The positivity threshold was as determined by the manufacturer (%S/N ≤ 40: positive result; 40 < %S/N ≤ 50: doubtful result; %S/N > 50: negative result). ELISA positive and doubtful samples, and the 9 samples that were not tested by ELISA, were then tested for the presence of neutralizing antibodies against both WNV and USUV using Virus Neutralizing Tests (VNT) as described in (Chevalier et al. 2025). The titer values of 0, 10, 20, 40, 80, 160 and 320 were then log2-transformed into respectively 0, 1, 2, 3, 4, 5 and 6. For model analysis, ELISA doubtful results were considered as positive, and ELISA sensitivity and specificity were estimated on that basis (see below).

### Model

#### Model description

In a context of WNV and USUV cocirculation, serological cross-reactivity complicates the interpretation of serological test results, even when they are considered highly specific like VNT. Diagnostic laboratories may therefore apply decision algorithms based on thresholds for VNT titers, e.g. attributing the WNV-positive serostatus to a sample if its WNV titer is ≥10 and at least four-fold that of other viruses’ titers (Chevalier et al. 2025) (here referred to as the “threshold method”, Supplementary Figure S1). Instead, here, we adopted a model-based approach to discriminate between WNV and USUV induced antibody responses (Hozé et al. 2021). Our model allowed to estimate (i) annual forces of infection (FOI) by WNV and USUV, and (ii) risk factors for infection, given ELISA and VNT serological test results and exposure data (time spent in the zoo and species category). We decomposed it into an epidemiological model, which inferred the true (unobserved) serological statuses for both viruses for each sample (*I*_*WNV*_ and *I*_*USU*_) using exposure variables (*X*), and two observation models associated to the Panflavivirus ELISA test results (*F*) and VNT results (*V*). The model likelihood can be written as:

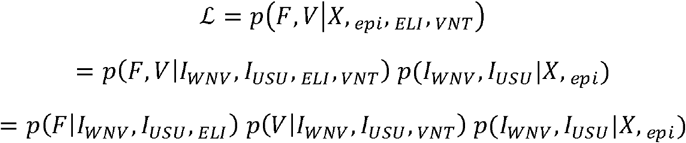

Where *θ*_*epi*_ represented the parameters of the epidemiological model, including viruses’ yearly FOI, the seroreversion rate (*τ*), the relative risk of infection in mammals and reptiles, and in birds with exposure to water (aquatic lifestyle and/or proximity to stagnant water in summer nights) as compared to birds with no water exposure (respectively *β*^*ma*^, *β*^*aqua*^, *β*^*stag*^ and *β*^*both*^) (Table 1). The parameters of the ELISA observation model (*θ*_*ELI*_) were the Panflavivirus ELISA sensitivity (*μ*_*pan*_, i.e. the probability of positive or doubtful ELISA test if the true serostatus is positive) and specificity (*γ*, i.e. the probability of negative ELISA test if the true serostatus is negative), assumed identical for both WNV and USUV. Finally, VNT observation model’s parameters (*θ*_*VNT*_) included the VNT sensitivity (*μ*_*VNT*_, assumed identical for both viruses), the mean titer values expected in respectively WNV (*α*_*WNV*_) and USUV (*α*_*USU*_) seropositive samples, and a parameter quantifying the VNT cross-reactivity between WNV and USUV (*ψ*) (Table 1). Full details on the model components are in Supplementary Note 1.

**Table 1.** Parameters of the epidemiological (Ө_epi_), ELISA observation (Ө_ELI_) and VNT observation (Ө_VNT_) models. Parameters were estimated in a Bayesian framework with either an uninformative or informative (based on the literature) prior distribution, or a fixed value based on the literature.

|  | Parameter | Description | Prior distribution or fixed value |
| --- | --- | --- | --- |
| *Epidemiological model - $\Theta_{\text{epi}}$* | $\tau$ | WNV and USUV annual seroreversion rate | 0.086<br>(Hamouche et al. 2025) |
| | $\lambda_{\text{WNV}, \leq 2020}$ | Pre-2020 annual force of infection by WNV (/year) | Exponential of Uniform(-20, 1) |
| | $f_{\text{WNV}, \geq 2021}$ | Relative annual force of infection by WNV in post-2021 years as compared to pre-2020 | Exponential of Uniform(-6, 10) |
| | $\lambda_{\text{WNV}, \geq 2021}$ | Annual force of infection by WNV in post-2021 years (/year) | $= f_{\text{WNV}, \geq 2021} \cdot \lambda_{\text{WNV}, \leq 2020}$ |
| | $\lambda_{\text{USUV}, \leq 2017}$ | Pre-2017 annual force of infection by USUV (/year) | Exponential of Uniform(-20, 1) |
| | $f_{\text{USUV}, y}$ | Relative annual force of infection by USUV in year y as compared to pre-2017 | Exponential of Uniform(-6, 10) |
| | $\lambda_{\text{USUV}, y}$ | Annual force of infection by USUV in year y (/year) | $= f_{\text{USUV}, y} \cdot \lambda_{\text{USUV}, \leq 2017}$ |
| | $\beta^{\text{ma}}$ | Relative risk of infection in mammals and reptiles, as compared to birds with no water exposure | Exponential of Uniform(-20, 3) |
| | $\beta^{\text{aqua}}$ | Relative risk of infection in birds with an aquatic lifestyle only, as compared to no water exposure | Exponential of Uniform(-20, 3) |
| | $\beta^{\text{stag}}$ | Relative risk of infection in birds exposed to stagnant water in summer nights only, as compared to no water exposure | Exponential of Uniform(-20, 3) |
| | $\beta^{\text{both}}$ | Relative risk of infection in birds with both an aquatic lifestyle and exposed to stagnant water in summer nights, as compared to no water exposure | Exponential of Uniform(-20, 3) |
| *ELISA model - $\Theta_{\text{ELI}}$* | $\mu_{\text{pan}}$ | Sensitivity of the Panflavivirus ELISA test for WNV and USUV | Beta(101.16, 3.90)<br>(Beck et al. 2017; Girl et al. 2024) |
| | $\gamma$ | Specificity of the Panflavivirus ELISA test for WNV and USUV | Beta(13.94, 1.17)<br>(Beck et al. 2017; Girl et al. 2024; Hogrefe et al. 2004) |
| *VNT model - $\Theta_{\text{VNT}}$* | $\varepsilon$ | Ratio of sensitivity of the VNT relatively to the ELISA test for WNV and USUV | Beta(6.76, 1.82) (Girl et al. 2024) |
| | $\mu_{\text{VNT}}$ | Sensitivity of the VNT for WNV and USUV | $= \varepsilon \cdot \mu_{\text{pan}}$ |
| | $\alpha_{\text{WNV}}$ | Expected (log2-transformed) VNT titer value in a sample from an individual previously infected with WNV | Uniform(3, 7) |
| | $\alpha_{\text{USUV}}$ | Expected (log2-transformed) VNT titer value in a sample from an individual previously infected with USUV | Uniform(3, 7) |
| | $\psi_{\text{WNV-USUV}}$ | Level of VNT cross-reactivity from WNV to USUV antibodies | Uniform(0.1, 0.6) |
| | $\psi_{\text{USUV-WNV}}$ | Level of VNT cross-reactivity from USUV to WNV antibodies | Uniform(0.1, 0.6) |

WNV: West Nile virus; USUV: Usutu virus; ELISA: Enzyme-Linked Immuno-Sorbent Assay; VNT: Virus Neutralization Test.

To clarify the notations, here, we designate with the terms “seropositivity”, “seroprevalence” and “serostatus” the true (unobserved but inferred) past infection by one and/or the other virus (*I*_*WNV*_ and *I*_*USU*_), while “ELISA positivity” and “VNT positivity” refer to the observed test results (*F* and *V*) that may differ because of imperfect test sensitivity or specificity.

In our model, we split estimates of WNV annual FOI in two periods, assuming different levels of viral circulation: pre-2020 (pre-emergence period) and 2021 onwards (post-emergence period). Concerning USUV, the surveillance of wild bird mortalities and human cases allowed to determine that the circulation was particularly high and widespread in France in 2018 as compared to previous years, and was also high in some of the following years (Duvignaud et al. 2024; Chevalier et al. 2025; Angeloni et al. 2023). This is why, for USUV, we estimated three annual FOI values: pre-2017, 2018 and 2019 onwards. For both pathogens, we tested alternative period splits as part of the sensitivity analysis (see below).

#### Model parameterization

The model was fitted to the data using a Bayesian Markov Chain Monte-Carlo (MCMC) algorithm implemented with the *rjags* package (Plummer et al. 2016) in R version 4.4.1. We assumed the VNT sensitivity to be a fraction (*ε*) of the Panflavivirus ELISA sensitivity, and we assumed the VNT specificity to be 1 (Hamouche et al. 2025; Beck et al. 2017). For some parameters (the seroreversion rate *τ*, the sensitivity *μ*_*pan*_ and specificity *γ* of the Panflavivirus ELISA test, and the ratio of sensitivity of the VNT relatively to the ELISA test *ε*), we used a fixed value or a prior distribution informed by previously published studies (see details in Supplementary Note 2 and Supplementary Figure S2). For the other parameters, we used uninformative priors (Table 1). In the main analysis, we assumed α_WNV_ = α_USU_ = α and *ψ*_*WNV-USU*_ = *ψ*_*USU-WNV*_ = *ψ*, and then performed a sensitivity analysis on these assumptions (see below).

Before fitting the model to the actual data, we validated the estimation framework with synthetic data. Indeed, we randomly drew 30 sets of mock (known) parameter values for the model and used them to simulate mock ELISA and VNT results for each serum in the dataset. We then fitted the model independently to each of the 30 synthetic datasets. This allowed to compare the known parameter values (that generated the synthetic data) to the parameter values estimated from the synthetic data.

#### Sensitivity analysis

We tested the robustness of our results to assumptions made as part of the model construction. We first relaxed the α_WNV_ = α_USU_ hypothesis, and then the *ψ*_*WNV-USU*_ = *ψ*_*USU-WNV*_ hypothesis, while keeping the other parameters identical. Moreover, we changed the splits in viral circulation periods assumed for estimating FOI. We fitted the model assuming four periods (pre-2017, 2018, 2019-2022 and post-2023) instead of three for USUV, assuming one period (with all years) or three (pre-2020, 2021 and post-2022) instead of two for WNV, or assuming two different periods for WNV (pre-2021 and post-2022 instead of pre-2020 and post-2021). We compared the Deviance Information Criterion (DIC) of these alternative models to the main model (Spiegelhalter et al. 2002).

## Results

### Serological results

Among 1707 serum samples collected in birds, mammals and reptiles of La Palmyre zoo between January 2015 and November 2024, 1698 were tested by Panflavivirus ELISA test and 430 (25.3%) were positive (Figure 1 and Supplementary Table S3). The proportion was 39.4% (407/1032), 3.50% (23/657) and 0% (0/9) in birds, mammals and reptiles, respectively.

When focusing on the 6 bird species with over 30 samples available, the proportion of ELISA positive tests by species ranged between 26.7% in *Phoenicopterus chilensis* and 88.6% in *Pelecanus onocrotalus* (see all results by species in Supplementary Table S3). Among 60 mammal species sampled (resp. 6 reptile species), 11 mammal species (resp. 0 reptile species) had at least one positive ELISA test: *Aonyx cinereus, Bison bison, Colobus guereza, Connochaetes taurinus, Elephas maximus, Hippopotamus amphibius, Macaca nemestrina, Mandrillus sphinx, Oryx dammah, Pan troglodytes* and *Ursus maritimus*.

Regarding the exposure of bird species to water, the proportion of ELISA positive samples was respectively 13.0% (3/23), 52.4% (118/225), 97.2% (35/36) and 33.6% (251/748) in birds with respectively no exposure to water, only an aquatic lifestyle, only a proximity to stagnant water in summer nights and both types of water exposure. By year of sampling, the proportion of positive ELISA tests varied between 0% (0/39) in 2015 and 35.8% (185/517) in 2023 (Supplementary Figure S3 and Supplementary Table S3).

A total of 430 samples that were Panflavivirus ELISA positive along with 9 samples that were not tested by ELISA were tested by VNT for WNV and USUV. Across interpretable results, 68.7% (298/434) of titer values were not null for WNV, 70.3% (300/427) for USUV, and 51.1% (216/423) for both viruses. Joint and marginal distributions of titer values observed for both viruses are shown in Figure 2.

**Figure 2.**
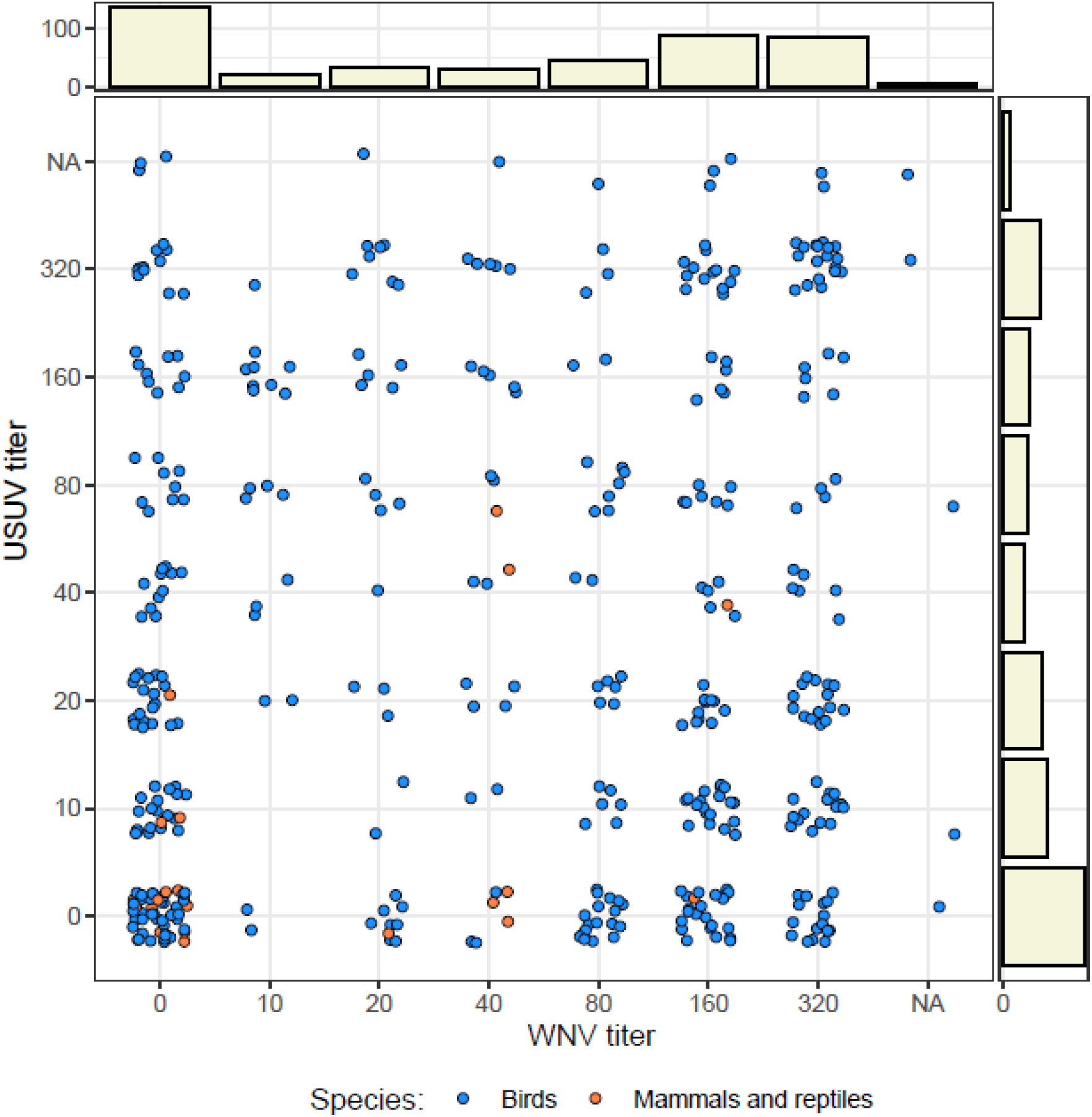
Titer results of the Virus Neutralization Tests for West Nile (WNV) and Usutu (USUV) viruses for 439 serological samples that were first positive to the Panflavivirus ELISA test (or untested), collected in birds, mammals and reptiles of La Palmyre zoo. Bar graphs represent the marginal distributions for each virus. “NA” values correspond to uninterpretable VNT titer results for one and/or the other virus.

The earliest sample presenting both a non-null WNV VNT titer (of 80) and an USUV VNT titer of 0 was in a Southern ground hornbill (*Bucorvus leadbeateri*) in July 2021. Nine birds sampled between June and November 2019 (7 *Balearica regulorum*, one *Alopochen aegyptiaca* and one *Phoenicopterus chilensis*) were the first individuals of the zoo with a non-null USUV VNT titer and a WNV VNT titer of 0.

### Model validation and goodness of fit

We built a model allowing to estimate the FOI by WNV and USUV across years in the zoo, risk factors for infection and the serological cross-reactivity between viruses during testing. When fitting the model to synthetic datasets, parameter estimates were consistent with the known parameters values that generated the data (Supplementary Figure S4). We then fitted the model to the actual data. The observed and predicted proportions of positive ELISA test results were consistent when stratified by year of sampling and by species category (Supplementary Figure S3). Moreover, the marginal and joint distributions of the observed vs. predicted titer values for WNV and USUV were concordant, suggesting a satisfying goodness of fit (Supplementary Figure S5).

### Estimated WNV and USUV forces of infection and risk factors

In the zoo, we estimated the annual rate of infection (FOI) by WNV in birds with no water exposure to be 0% (95% Highest Posterior Density Interval: [0%; 0%]) prior to 2020, and 6% [3%; 11%] in the period 2021-2024 (Figure 3). Regarding USUV in birds with no water exposure, the annual FOI was estimated at 0% [0%; 0%] for the pre-2017 period, at 8% [3%; 14%] in 2018, and at 1% [0%; 2%] for the period 2019-2024 (Figure 3 and Supplementary Figure S6).

**Figure 3.**
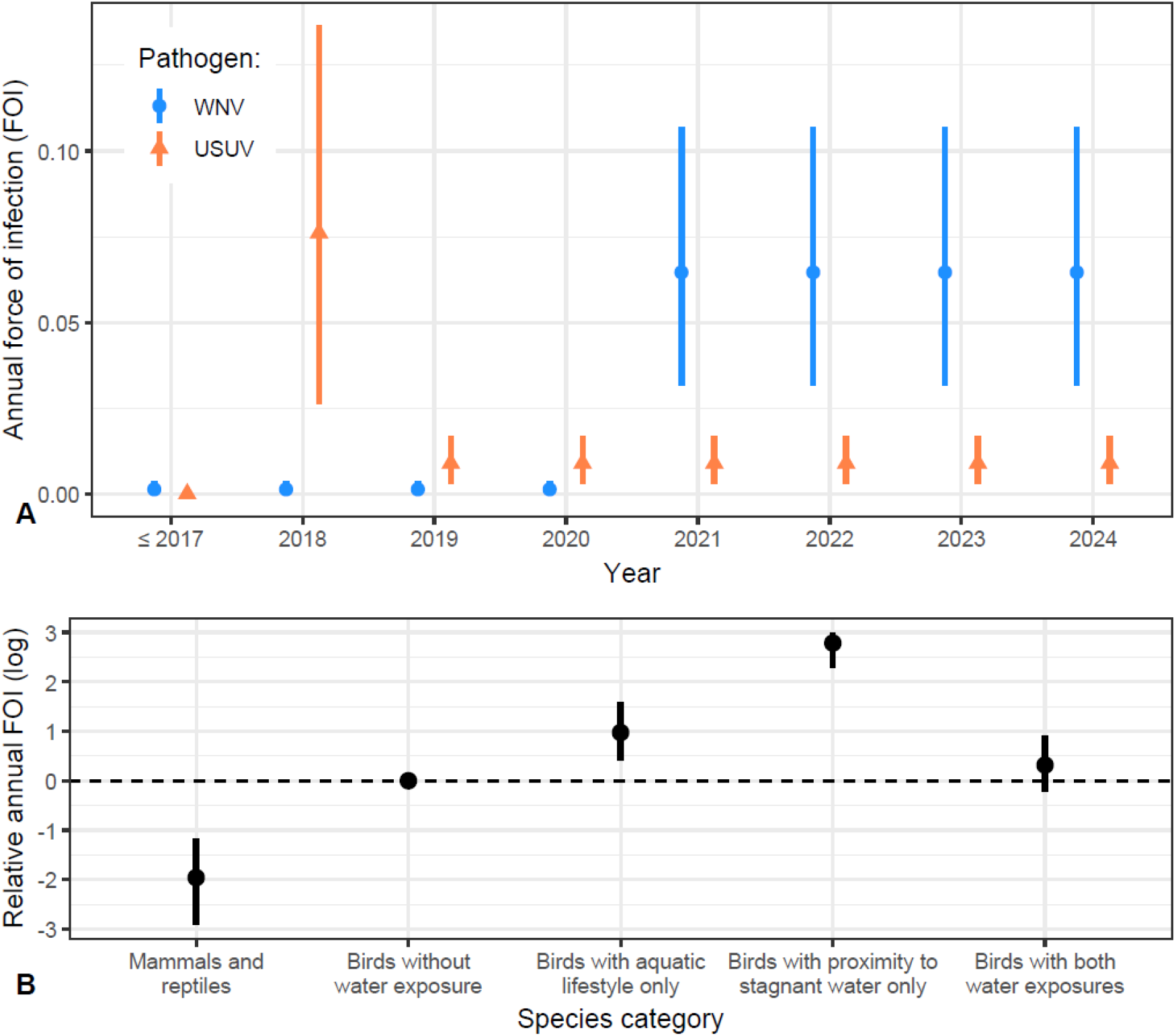
Panel A: Posterior estimates (median and 95% credible interval) of Λ_k,y(t)_, the annual forces of infection (FOI) in birds with no water exposure, for West Nile (WNV, blue circles) and Usutu (USUV, orange triangles) viruses in La Palmyre zoo. We considered the exposure time was only between May 1^st^ and November 30^th^ of each year (the mosquito season). Panel B: Posterior estimates (median and 95% credible interval) of the log-scaled relative risk of infection in mammals and reptiles, and in birds with exposure to water (aquatic lifestyle and/or proximity to stagnant water in summer nights) as compared to birds with no water exposure (respectively β^ma^, β^aqua^, β^stag^ and β^both^).

For both viruses, the relative risk of infection in mammals and reptiles was around seven times lower than in birds with no water exposure (0.14 [0.05; 0.28]). It was 2.7 [1.3; 4.4], 16.2 [9.9; 20.1] and 1.4 [0.7; 2.3] in birds with only an aquatic lifestyle, only a proximity to stagnant water in summer nights and both types of water exposure respectively (Figure 3 and Supplementary Figures S6 and S7). Correlations between parameters are shown in Supplementary Figure S8.

Based on the fitted model, we inferred for each year and each virus (i) the true seroprevalence (i.e. the predicted proportion of samples with a positive true serostatus), and (ii) the predicted probability that at least one of the animals sampled at the zoo (that year or before) had been infected, as depicted in Figure 4. Regarding the latter probability for WNV, the 95% credible interval was close to 0 until 2020, and then increased to 0.86 [0.30; 0.99] in 2021 and 0.99 [0.99; 1.0] in 2022 (Figure 4). Moreover, the probability that USUV infected at least one animal previously sampled at the zoo increased from 0.0 [0.0; 0.05] in 2017 to 0.21 [0.10; 0.35] in 2018 and 0.99 [0.99; 1.0] in 2019. The estimated true seroprevalence in sampled animals increased from 0% [0%; 0%] for both WNV and USUV in 2015 to 28% [25%; 31%] for WNV and 13% [11%; 15%] for USUV in 2024 (Figure 4).

**Figure 4.**
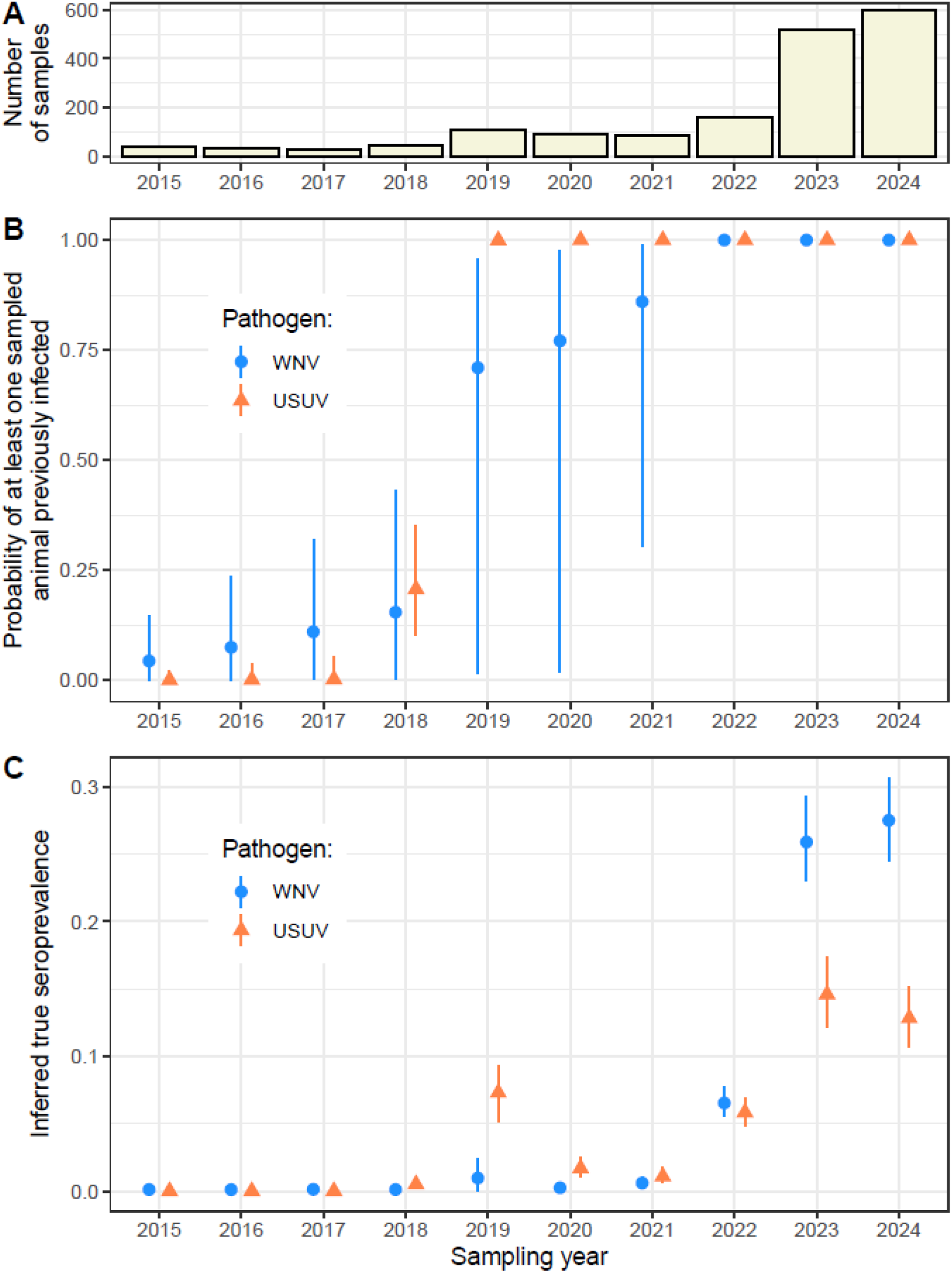
For each year, number of samples collected at the zoo (panel A), estimated probability of having at least one sampled animal that was truly seropositive to each virus until that year (panel B), and estimated true seroprevalence for each virus (panel C). Points and intervals are respectively the median and 95% credible interval of the posterior distributions. The latter can be too small to be visible in panels B and C of the figure.

### Estimated performance and cross-reactivity of the serological tests

Although estimated with informative priors, the median posterior value of the sensitivity and specificity of the Panflavivirus ELISA test for the presence of WNV or USUV antibodies were 96.1% [91.9%; 99.1%] and 99.1% [98.6%; 99.6%] respectively (Table 2 and Supplementary Figure S6). The VNT sensitivity estimate was 80.4% [76.5%; 84.3%]. In case of detection by the VNT, the expected (log2-transformed) titer response in a serum from an individual previously infected by any of the two viruses was estimated at 4.7 [4.5; 4.9]. Moreover, we estimated that, if an individual had been infected by a first virus, the log2-tranformed titer value expected during the VNT for the second virus increased by a fraction of 0.34 [0.29; 0.38] of the titer value expected for the first virus (Table 2 and Supplementary Figure S6).

**Table 2.** Estimates (median and 95% Highest Posterior Density Interval) of the sensitivity and specificity of ELISA and VNT tests, VNT expected titer values and serological cross-reactivity (Ө_ELI_ and Ө_VNT_ observation model parameters).

| Parameter | Description | Median [95% credible interval] of the posterior distribution |
| --- | --- | --- |
| $\mu_{\text{pan}}$ | Sensitivity of the Panflavivirus ELISA test for WNV and USUV | 96.1% [91.9%; 99.1%] |
| $\gamma$ | Specificity of the Panflavivirus ELISA test for WNV and USUV | 99.1% [98.6%; 99.6%] |
| $\mu_{\text{VNT}}$ | Sensitivity of the VNT for WNV and USUV | 80.4% [76.5%; 84.3%] |
| $\alpha$ | Expected (log2-transformed) VNT titer value in a sample from an individual previously infected with WNV (or USUV) | 4.7 [4.5; 4.9] |
| $\psi$ | Level of VNT cross-reactivity between WNV and USUV antibodies | 0.34 [0.29; 0.38] |

ELISA: Enzyme-Linked Immuno-Sorbent Assay; VNT: Virus Neutralization Test.

Using these estimates, we also computed a model-based probability for a sample to be seropositive to WNV and USUV, given values of the titer for both viruses (independently of the sampled individual’s characteristics), as detailed in the Supplementary Note 3. This model-based classification can be compared with the threshold method, more classically used in the lab for the distinction between WNV and USUV, depicted in Supplementary Figure S1. From our model, we found that the probability of seropositivity to each virus strongly depended on the VNT titer value for both viruses (Figure 5). Therefore, in such a context of co-circulation, VNT should be performed for both cross-reacting viruses in order to be able to interpret the titer value. Furthermore, this model-based classification was sensitive to the true seroprevalence of both viruses in the sample (Supplementary Figure S9), which we inferred from the data in our study but which could be informed otherwise in the future (e.g. using data from an area where a virus is known to be absent).

**Figure 5.**
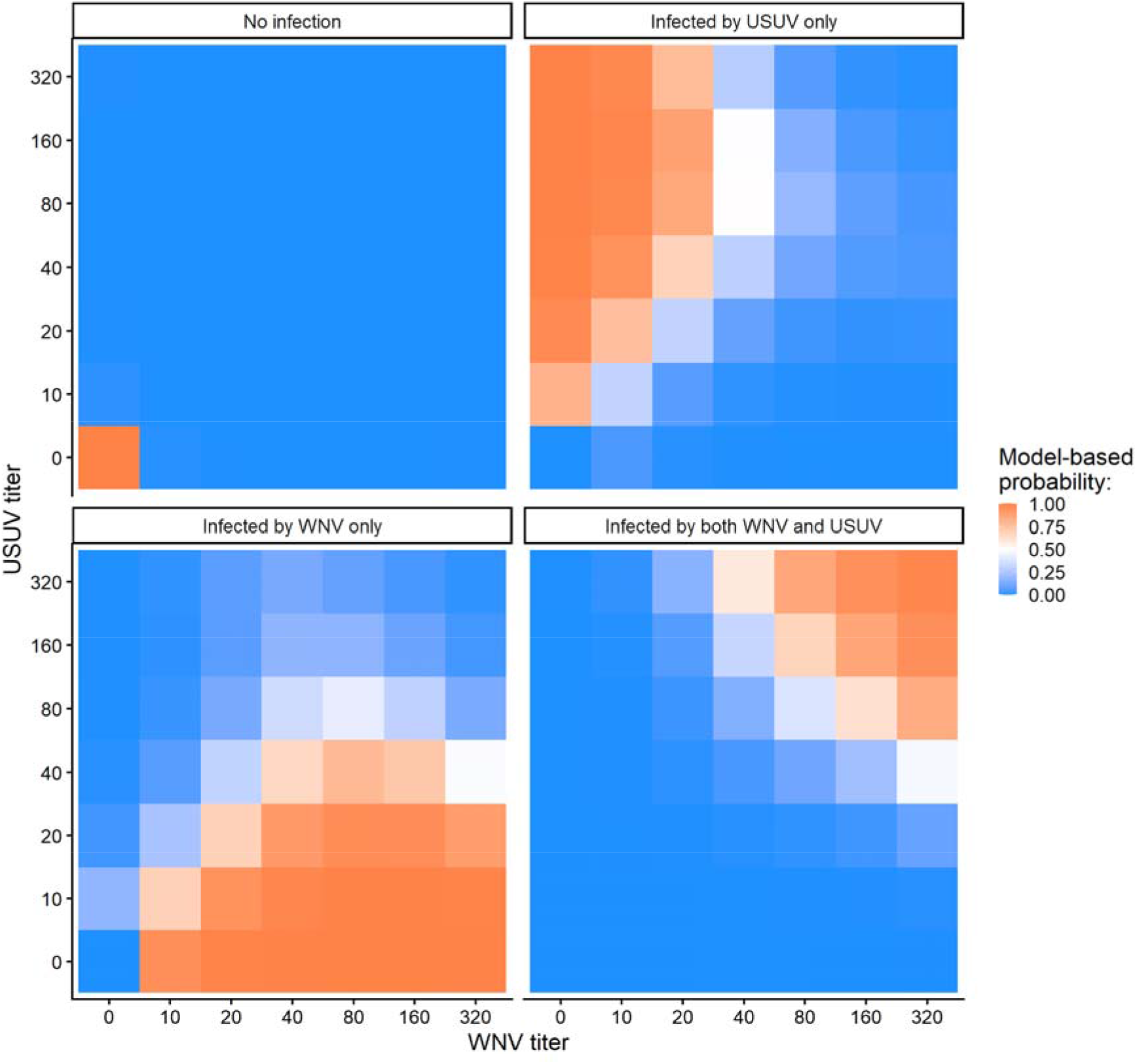
Model-based probability of true serological status (past infection) of a sample to USUV and/or WNN, given VNT titer values for both viruses and using the global true seroprevalences (i.e. the proportion of positive true serostatus) inferred for the whole dataset (see details on the computation in Supplementary Note 3).

### Sensitivity analysis

All alternative models had a higher DIC than the main model, except for the model with four instead of three circulation periods for USUV (Supplementary Table S4). However, the mixing of chains during MCMC was less good for this latter model, and the posterior estimates of the 2019-2022 and 2023-2024 USUV FOIs were similar (Supplementary Figure S10). This is why we did not favor this alternative model to the main model.

## Discussion

In this study, we analyzed serological data obtained from zoo animals within a modelling framework to unravel the dynamics of two cross-reacting zoonotic vector-borne viruses in an emerging area. Our findings have implications for both the epidemiology and the testing of these orthoflaviviruses.

First, we found that WNV was already spreading unnoticed in the zoo possibly as early as 2021, even though the first case reported by the animal and human passive surveillance systems in the region was in 2022 at a 70 km distance (Chevalier et al. 2025). It confirms that WNV is able to spread silently (Fritsch et al. 2022; Hamouche et al. 2025), because (i) infection symptoms may not be systematic nor specific to this virus, or because (ii) morbidity and mortality events may go unnoticed, depending on the population affected and the levels of circulation. Our results also support that animals in zoos can help to identify temporal patterns of zoonotic vector-borne diseases emergence, as they ensure the strict health monitoring of animals of diverse taxa and are often close to human dwellings (Hernandez-Colina et al. 2024; Van Leeuwen et al. 2023). This may especially be the case when alternative surveillance systems present a low sensitivity, for instance due to the low density of other animal populations (e.g. few equids in cities to monitor WNV). In practice, samples could be collected regularly in sentinel zoos, from animals or in zoo waterbodies (McGregor et al. 2023; Adesola et al. 2025). Regarding WNV and USUV specifically, our results suggest that sampling birds with a high exposure to water (e.g. flamingos), or their environment, would be most efficient, although target animals may be different for other pathogens. In the future, it would be interesting to also evaluate the costs associated with different screening strategies.

Using all the data at once, our model associated measured antibody responses to the probability of infection by one or the other virus, accounting for serological cross-reactivity. As compared to the more classical threshold method that attributes the causative pathogen based on threshold rules applied to VNT titer values (see Supplementary Figure S1), our method allows (i) to provide a measure of uncertainty of the serostatus, and (ii) to make the distinction between samples from coinfected animals and samples with uncertain serostatus. In the future, such model-informed diagnosis may assist routine surveillance activities, in a context of growing geographical overlap between vector-borne diseases that have similar symptomatology and that sometimes present serological cross-reactivity (Lustig et al. 2018; Madere et al. 2025). To illustrate this point, our model-based approach allowed to attribute a probability of 0.86 [0.30; 0.99] to the assertion that WNV was circulating in the zoo prior to the summer of 2021.

In our study, birds were more infected by WNV and USUV than mammals and reptiles, consistently with previous studies (Chen et al. 2024; Ludwig et al. 2002). Birds are indeed recognized as primary hosts for both viruses (Simonin 2024) and *Culex pipiens*, which may be their main vector in the region (Bigeard et al. 2024; Duvignaud et al. 2024), tends to be ornithophilic (Bobeva et al. 2025; Tiron et al. 2021; Muñoz et al. 2012). While WNV virulence varies across host species (Komar et al. 2005), the virus was identified as responsible for an outbreak that affected the zoo’s flamingo population in 2023 (Duvignaud et al. 2024). The reason might be that flamingos are more exposed to mosquito vectors and/or to the orofecal transmission route, as they feed by filtering pond water with their bill (Delfino and Carlos 2022).

Indeed, besides the taxon-related risk factor, our results show a positive effect of two types of birds’ exposure to waterbodies on the infection risk. First, the effect found for bird species with an aquatic lifestyle is consistent with a higher exposure of individuals to the orofecal route, which has been proposed as a possible transmission route for both WNV and USUV (Hinton et al. 2015; Benzarti et al. 2020). Second, the effect of birds’ exposure to stagnant water may be explained by a higher abundance of *Culex* vectors around water ponds at local scale, especially if the water is rich in organic matter (Wouters et al. 2024; Pautzke et al. 2024), although these insects are known to be able to fly across several hundred meters (Verdonschot and Besse-Lototskaya 2014). At the landscape scale, the presence of stagnant water (or its proxies) has been suggested to favor WNV spread by impacting mosquito breeding and/or bird-mosquito interactions, even though this relationship depends on the season, region and landcover (Esser et al. 2019). In (Roche et al. 2015), the “Wetland habitat use” ecologic trait was not associated with WNV seroprevalence in bird species. Because our results were obtained from a small zoo area, they may not be generalizable to the larger ecosystem or global scale.

Even though we found USUV circulated in the settings every year from 2018 to 2024, no important mortality event associated with this virus was noticed in this zoo’s animals. This may be due to a lower virulence of USUV as compared to WNV (Marshall et al. 2025), although the former has also been reported to cause mortality in some species of birds (e.g. *Strix nebulosa* and *Turdus merula*) and, sometimes, symptomatic infections in mammals, including humans (Chen et al. 2024; Angeloni et al. 2023). In a previous zoo study led in an endemic area for both viruses (Constant et al. 2020), USUV seroprevalence in birds was ten times higher than that of WNV. Here, the model-derived seroprevalence was higher for WNV than for USUV starting from 2023, due to its estimated higher force of infection in an emergence context. In the future, the dynamics of co-circulation of these two closely related orthoflaviviruses in the wild bird reservoir, accounting for (partial) cross-immunity and different introduction times, could be explored using a mechanistic model (Nikolay 2015).

Our analysis framework was based on modelling assumptions. First, we considered that the performances of both the Panflavivirus ELISA test and VNT were similar in birds than in other species, whereas antibody response or detection may vary across species (Fassbinder-Orth et al. 2016). Nevertheless, in this study, we used competition ELISA tests that have been validated on multiple species as specified by the manufacturer. Second, we did not make the distinction between neutralizing and non-neutralizing antibodies in the model, but instead assumed that the VNT sensitivity was lower than the ELISA sensitivity, precisely because the former does not detect non-neutralizing antibodies (Rabenau et al. 2007). Third, our estimates depend on the epidemiological hypotheses on viral circulation periods for WNV and USUV, and on the data used to fit the model. In that regard, depending on the epidemiological situation (e.g. a sample collected from an area where one of the viruses is expected to be absent), our model-based approach of diagnosis can result in different interpretations of a same set of VNT results. Therefore, expanding our model to zoos in multiple areas would require to account for this diversity of epidemiological backgrounds, besides collecting the same set of exposure data (time of presence at the zoo and species category).

To conclude, our modelling framework fitted with serological data collected in zoo animals allowed to reconstruct the timing of emergence of WNV in the area, despite its cross-reactivity with USUV. It highlights the utility of zoo settings in assessing zoonotic vector-borne diseases dynamics. Such approaches mixing quantitative methods to serum bank databases will be key to improve surveillance in a future with growing arboviruses burden.

## Data Availability

All data produced in the present study are available upon reasonable request to the authors.

## Acknowledgements

We thank the staff of La Palmyre Zoo for their assistance during the sampling campaigns conducted in 2023 and 2024 alongside the influenza vaccination program. This work was supported by a DIM1Health postdoctoral fellowship awarded by the Conseil Régional d’Ile-de-France, in addition to support from LabEx IBEID and the ANRS-MIE Emergence program. The funders had no role in study design, data collection and analysis, decision to publish or preparation of the manuscript.

## Ethics statement

Blood sampling was opportunistic and performed for a primary purpose unrelated to the study. Samples were collected during routine veterinary procedures (e.g., influenza vaccination). The protocol was approved by VetAgro-Sup’s ethics committee (agreement number 2490).

## Bibliography

Adesola, Ridwan Olamilekan, Adetolase Azizat Bakre, Bamidele Nyemike Ogunro, et al. 2025. “Avian Influenza Screening in Captive Wild Birds and Biosecurity Appraisal of Zoological Gardens in Southwestern Nigeria.” Veterinary Medicine International 2025 (1): 3419266. 10.1155/vmi/3419266.

Angeloni, Giorgia, Michela Bertola, Elena Lazzaro, et al. 2023. “Epidemiology, Surveillance and Diagnosis of Usutu Virus Infection in the EU/EEA, 2012 to 2021.” Eurosurveillance 28 (33): 2200929. 10.2807/1560-7917.ES.2023.28.33.2200929.

Banet-Noach, Caroline, Lubov Simanov, and Mertyn Malkinson. 2003. “Direct (Non-Vector) Transmission of West Nile Virus in Geese.” Avian Pathology: Journal of the W.V.P.A 32 (5): 489–94. 10.1080/0307945031000154080.

Beck, Cécile, Steeve Lowenski, Benoit Durand, Céline Bahuon, Stéphan Zientara, and Sylvie Lecollinet. 2017. “Improved Reliability of Serological Tools for the Diagnosis of West Nile Fever in Horses within Europe.” PLOS Neglected Tropical Diseases 11 (9): e0005936. 10.1371/journal.pntd.0005936.

Bellegarde de Saint Lary, Chiara de, Louella M. R. Kasbergen, Patricia C. J. L. Bruijning-Verhagen, et al. 2023. “Assessing West Nile Virus (WNV) and Usutu Virus (USUV) Exposure in Bird Ringers in the Netherlands: A High-Risk Group for WNV and USUV Infection?” One Health (Amsterdam, Netherlands) (Netherlands) 16 (June): 100533. 10.1016/j.onehlt.2023.100533.

Benzarti, Emna, José Rivas Michaël Sarlet, et al. 2020. “Experimental Usutu Virus Infection in Domestic Canaries Serinus Canaria.” Viruses 12 (2): 2. 10.3390/v12020164.

Bigeard, Clément, Laura Pezzi, Raphaelle Klitting, et al. 2024. “Molecular Xenomonitoring (MX) Allows Real-Time Surveillance of West Nile and Usutu Virus in Mosquito Populations.” PLOS Neglected Tropical Diseases 18 (12): e0012754. 10.1371/journal.pntd.0012754.

Bobeva, Aneliya, Martin P. Marinov, Stefania Klayn, Mihaela Ilieva, Dimitar Dimitrov, and Pavel Zehtindjiev. 2025. “Feeding Preferences of Mosquitoes (Culicidae) from the Eastern Balkans and Their Role in Transmission of Avian Malaria.” Integrative Zoology, ahead of print, November 11. 10.1111/1749-4877.70018.

Caballero-Gómez, J., D. Cano-Terriza, S. Lecollinet, et al. 2020. “Evidence of Exposure to Zoonotic Flaviviruses in Zoo Mammals in Spain and Their Potential Role as Sentinel Species.” Veterinary Microbiology 247 (August): 108763. 10.1016/j.vetmic.2020.108763.

Cabanová, Viktória, Jana Kerlik, Peter Kirschner, et al. 2023. “Co-Circulation of West Nile, Usutu, and Tick-Borne Encephalitis Viruses in the Same Area: A Great Challenge for Diagnostic and Blood and Organ Safety.” Viruses 15 (2): 366. 10.3390/v15020366.

Cano-Terriza, David, Rafael Guerra, Sylvie Lecollinet, et al. 2015. “Epidemiological Survey of Zoonotic Pathogens in Feral Pigeons (Columba Livia Var. Domestica) and Sympatric Zoo Species in Southern Spain.” Comparative Immunology, Microbiology and Infectious Diseases 43 (December): 22–27. 10.1016/j.cimid.2015.10.003.

Che-Castaldo, Judy P., Amy Byrne, Kaitlyn Perišin, and Lisa J. Faust. 2019. “Sex-Specific Median Life Expectancies from Ex Situ Populations for 330 Animal Species.” Scientific Data 6 (1): 1. 10.1038/sdata.2019.19.

Chen, Jiahao, Yuanyuan Zhang, Xiaoai Zhang, et al. 2024. “Epidemiology and Ecology of Usutu Virus Infection and Its Global Risk Distribution.” Viruses 16 (10): 1606. 10.3390/v16101606.

Chevalier, Noémie, Camille V. Migné, Teheipuaura Mariteragi-Helle, et al. 2025. “Seroprevalence of West Nile, Usutu and Tick-Borne Encephalitis Viruses in Equids from South-Western France in 2023.” Veterinary Research 56 (1): 1. 10.1186/s13567-025-01508-w.

Constant, Orianne, Karine Bollore, Marion Clé, et al. 2020. “Evidence of Exposure to USUV and WNV in Zoo Animals in France.” Pathogens 9 (12): 12. 10.3390/pathogens9121005.

Delfino, Henrique Cardoso, and Caio J. Carlos. 2022. “What Do We Know about Flamingo Behaviors? A Systematic Review of the Ethological Research on the Phoenicopteridae (1978– 2020).” Acta Ethologica 25 (1): 1–14. 10.1007/s10211-021-00381-y.

Duvignaud, A., C. Bigeard, A. Fontaine, et al. 2024. “Investigation One Health de l’émergence Des Virus West Nile et Usutu En Nouvelle Aquitaine.” Médecine et Maladies Infectieuses Formation, 24es Journées Nationales d’Infectiologie, vol. 3 (2, Supplement): S9. 10.1016/j.mmifmc.2024.04.018.

ECDC. 2025. “Historical Data by Year - West Nile Virus Seasonal Surveillance.” European Centre for Disease Prevention and Control. https://www.ecdc.europa.eu/en/west-nile-fever/surveillance-and-disease-data/historical.

Esser, Helen J., Ramona Mögling, Natalie B. Cleton, et al. 2019. “Risk Factors Associated with Sustained Circulation of Six Zoonotic Arboviruses: A Systematic Review for Selection of Surveillance Sites in Non-Endemic Areas.” Parasites & Vectors 12 (1): 1. 10.1186/s13071-019-3515-7.

Farfán-Ale José A., Bradley J. Blitvich, Nicole L. Marlenee, et al. 2006. “Antibodies to West Nile Virus in Asymptomatic Mammals, Birds, and Reptiles in the Yucatan Peninsula of Mexico.” The American Journal of Tropical Medicine and Hygiene 74 (5): 908–14.

Fassbinder-Orth, Carol A., Travis E. Wilcoxen, Tiffany Tran, et al. 2016. “Immunoglobulin Detection in Wild Birds: Effectiveness of Three Secondary Anti-Avian IgY Antibodies in Direct ELISAs in 41 Avian Species.” Methods in Ecology and Evolution 7 (10): 1174–81. 10.1111/2041-210X.12583.

Fritsch, Hegger, Felicidade Mota Pereira, Erica Azevedo Costa, et al. 2022. “Retrospective Investigation in Horses with Encephalitis Reveals Unnoticed Circulation of West Nile Virus in Brazil.” Viruses 14 (7). 10.3390/v14071540.

Girl, Philipp, Kathrin Euringer, Mircea Coroian, Andrei Daniel Mihalca, Johannes P. Borde, and Gerhard Dobler. 2024. “Comparison of Five Serological Methods for the Detection of West Nile Virus Antibodies.” Viruses 16 (5): 5. 10.3390/v16050788.

Hamouche, Celia, Jennifer Pradel, Nonito Pagès, et al. 2025. “Reconstructing the Silent Circulation of West Nile Virus in a Caribbean Island during 15 Years Using Sentinel Serological Data.” PLOS Neglected Tropical Diseases 19 (6): e0012895. 10.1371/journal.pntd.0012895.

Hartemink, N. A., S. A. Davis, P. Reiter, Z. Hubálek, and J. A. P. Heesterbeek. 2007. “Importance of Bird-to-Bird Transmission for the Establishment of West Nile Virus.” Vector-Borne and Zoonotic Diseases 7 (4): 575–84. 10.1089/vbz.2006.0613.

Hernandez-Colina, Arturo, Nicola Seechurn, Taiana Costa, Javier Lopez, Matthew Baylis, and Jenny C. Hesson. 2024. “Surveillance of Culex Spp. Vectors and Zoonotic Arboviruses at a Zoo in the United Kingdom.” Heliyon 10 (4): e26477. 10.1016/j.heliyon.2024.e26477.

Hinton, M. G., W. K. Reisen, S. S. Wheeler, and A. K. Townsend. 2015. “West Nile Virus Activity in a Winter Roost of American Crows (Corvus Brachyrhynchos): Is Bird-To-Bird Transmission Important in Persistence and Amplification?” Journal of Medical Entomology 52 (4): 683–92. 10.1093/jme/tjv040.

Hogrefe, Wayne R., Ronald Moore, Mary Lape-Nixon, Michael Wagner, and Harry E. Prince. 2004. “Performance of Immunoglobulin G (IgG) and IgM Enzyme-Linked Immunosorbent Assays Using a West Nile Virus Recombinant Antigen (preM/E) for Detection of West Nile Virus- and Other Flavivirus-Specific Antibodies.” Journal of Clinical Microbiology 42 (10): 4641– 48. 10.1128/JCM.42.10.4641-4648.2004.

Hozé, Nathanaël, Issa Diarra, Abdoul Karim Sangaré, et al. 2021. “Model-Based Assessment of Chikungunya and O’nyong-Nyong Virus Circulation in Mali in a Serological Cross-Reactivity Context.” Nature Communications 12 (1): 6735. 10.1038/s41467-021-26707-9.

Hozé, Nathanaël, Henrik Salje, Dominique Rousset, et al. 2020. “Reconstructing Mayaro Virus Circulation in French Guiana Shows Frequent Spillovers.” Nature Communications 11 (1): 2842. 10.1038/s41467-020-16516-x.

Komar, Nicholas, Nicholas A. Panella, Stanley A. Langevin, et al. 2005. “Avian Hosts for West Nile Virus in St. Tammany Parish, Louisiana, 2002.” American Journal of Tropical Medicine and Hygiene 73 (6): 1031.

Kuchinsky, Sarah C., Francesca Frere, Nora Heitzman-Breen, et al. 2021. “Pathogenesis and Shedding of Usutu Virus in Juvenile Chickens.” Emerging Microbes & Infections 10 (1): 725–38. 10.1080/22221751.2021.1908850.

Kvapil, Pavel, Joško Račnik, Marjan Kastelic, et al. 2021. “A Sentinel Serological Study in Selected Zoo Animals to Assess Early Detection of West Nile and Usutu Virus Circulation in Slovenia.” Viruses 13 (4): 4. 10.3390/v13040626.

Lecollinet, Sylvie, Yannick Blanchard, Christine Manson, et al. 2016. “Dual Emergence of Usutu Virus in Common Blackbirds, Eastern France, 2015.” Emerging Infectious Diseases 22 (12): 2225–27. 10.3201/eid2212.161272.

Ludwig, George V., Paul P. Calle, Joseph A. Mangiafico, et al. 2002. An Outbreak of West Nile Virus in a New York City Captive Wildlife Population. The American Journal of Tropical Medicine and Hygiene. July 1. 10.4269/ajtmh.2002.67.67.

Lustig, Yaniv, Danit Sofer, Efrat Dahan Bucris, and Ella Mendelson. 2018. “Surveillance and Diagnosis of West Nile Virus in the Face of Flavivirus Cross-Reactivity.” Frontiers in Microbiology 9 (October). 10.3389/fmicb.2018.02421.

Madere, Ferralita S., Aurea Virginia Andrade da Silva, Efemena Okeze, et al. 2025. “Flavivirus Infections and Diagnostic Challenges for Dengue, West Nile and Zika Viruses.” Npj Viruses 3 (1): 36. 10.1038/s44298-025-00114-z.

Marshall, Eleanor M., Luisa Barzon, Marion Koopmans, and Barry Rockx. 2025. “Extrapolation and Comparison of West Nile Virus- and Usutu Virus-Associated Neurological Diseases in Humans: Linking Pathology to Clinical Symptoms.” Clinical Microbiology Reviews 38 (3): e00232–24. 10.1128/cmr.00232-24.

McGregor, Bethany L., Lindsey M. Reister-Hendricks, Cale Nordmeyer, et al. 2023. “Using Zoos as Sentinels for Re-Emerging Arboviruses: Vector Surveillance during an Outbreak of Epizootic Hemorrhagic Disease at the Minnesota Zoo.” Pathogens 12 (1). 10.3390/pathogens12010140.

Muñoz, Joaquín, Santiago Ruiz, Ramón Soriguer, et al. 2012. “Feeding Patterns of Potential West Nile Virus Vectors in South-West Spain.” PLOS ONE 7 (6): e39549. 10.1371/journal.pone.0039549.

Neo, Jacqueline Pei Shan, and Boon Huan Tan. 2017. “The Use of Animals as a Surveillance Tool for Monitoring Environmental Health Hazards, Human Health Hazards and Bioterrorism.” Veterinary Microbiology 203 (May): 40–48. 10.1016/j.vetmic.2017.02.007.

Nikolay, Birgit. 2015. “A Review of West Nile and Usutu Virus Co-Circulation in Europe: How Much Do Transmission Cycles Overlap?” Transactions of The Royal Society of Tropical Medicine and Hygiene 109 (10): 609–18. 10.1093/trstmh/trv066.

O’Driscoll, Megan, Nathanael Hoze, Noemie Lefrancq, et al. 2024. “Epidemiological Inferences from Serological Responses to Cross-Reacting Pathogens.” medRxiv, 2024.08. 12.24311852.

Pautzke, Kellen C., Allan S. Felsot, John P. Reganold, and Jeb P. Owen. 2024. “Effects of Soil on the Development, Survival, and Oviposition of Culex Quinquefasciatus (Diptera: Culicidae) Mosquitoes.” Parasites & Vectors 17 (1): 154. 10.1186/s13071-024-06202-y.

Plummer, Martyn, Alexey Stukalov, Matt Denwood, and Maintainer Martyn Plummer. 2016. “Package ‘Rjags.’” Vienna, Austria.

Rabenau, Holger F., Branko Marianov, Sabine Wicker, and Regina Allwinn. 2007. “Comparison of the Neutralizing and ELISA Antibody Titres to Measles Virus in Human Sera and in Gamma Globulin Preparations.” Medical Microbiology and Immunology 196 (3): 151–55. 10.1007/s00430-007-0037-2.

Roche, Benjamin, Serge Morand, Eric Elguero, Thomas Balenghien, Jean-François Guégan, and Nicolas Gaidet. 2015. “Does Host Receptivity or Host Exposure Drives Dynamics of Infectious Diseases? The Case of West Nile Virus in Wild Birds.” Infection, Genetics and Evolution 33 (July): 11–19. 10.1016/j.meegid.2015.04.011.

Simonin, Yannick. 2024. “Circulation of West Nile Virus and Usutu Virus in Europe: Overview and Challenges.” Viruses 16 (4): 4. 10.3390/v16040599.

Spiegelhalter, David J., Nicola G. Best, Bradley P. Carlin, and Angelika Van Der Linde. 2002. “Bayesian Measures of Model Complexity and Fit.” Journal of the Royal Statistical Society: Series B (Statistical Methodology) 64 (4): 4.

Tiron, Georgiana Victoria, Ioana Georgeta Stancu, Sorin Dinu, et al. 2021. “Characterization and Host-Feeding Patterns of Culex Pipiens s.l. Taxa in a West Nile Virus-Endemic Area in Southeastern Romania.” Vector-Borne and Zoonotic Diseases 21 (9): 713–19. 10.1089/vbz.2020.2739.

Van Leeuwen, Pauline, Sarah Falconer, Jasmine Veitch, et al. 2023. “Zoos as Sentinels? A Meta-Analysis of Seroprevalence of Terrestrial Mammalian Viruses in Zoos.” EcoHealth 20 (1): 43–52. 10.1007/s10393-023-01635-w.

Verdonschot, Piet F. M., and Anna A. Besse-Lototskaya. 2014. “Flight Distance of Mosquitoes (Culicidae): A Metadata Analysis to Support the Management of Barrier Zones around Rewetted and Newly Constructed Wetlands.” Limnologica 45 (March): 69–79. 10.1016/j.limno.2013.11.002.

Veronesi, Rodolfo, Gregorio Gentile, Marco Carrieri, Bettina Maccagnani, Luisa Stermieri, and Romeo Bellini. 2012. “Seasonal Pattern of Daily Activity of Aedes Caspius, Aedes Detritus, Culex Modestus, and Culex Pipiens in the Po Delta of Northern Italy and Significance for Vector-Borne Disease Risk Assessment.” Journal of Vector Ecology 37 (1): 49–61. 10.1111/j.1948-7134.2012.00199.x.

Wouters, Roel M., Wouter Beukema, Maarten Schrama, et al. 2024. “Local Environmental Factors Drive Distributions of Ecologically-Contrasting Mosquito Species (Diptera: Culicidae).” Scientific Reports 14 (1): 19315. 10.1038/s41598-024-64948-y.

