## Supplementary materials for "Modelling serological cross-reactivity to disentangle the dynamics of West Nile and Usutu viruses in an emerging area"

**Supplementary Table S1** (provided as a separate file). Description of the animal species sampled in the study: number of samples, number of individuals sampled, first and last year of sampling, species category (including the level of exposure to waterbodies within the zoo).

**Supplementary Table S2** (provided as a separate file). Number of animals kept at the zoo by species, as of February 2020 (median month of our study).

**Supplementary Table S3** (provided as a separate file). Panflavivirus ELISA results observed for zoo animals sampled in the study, overall and by category.

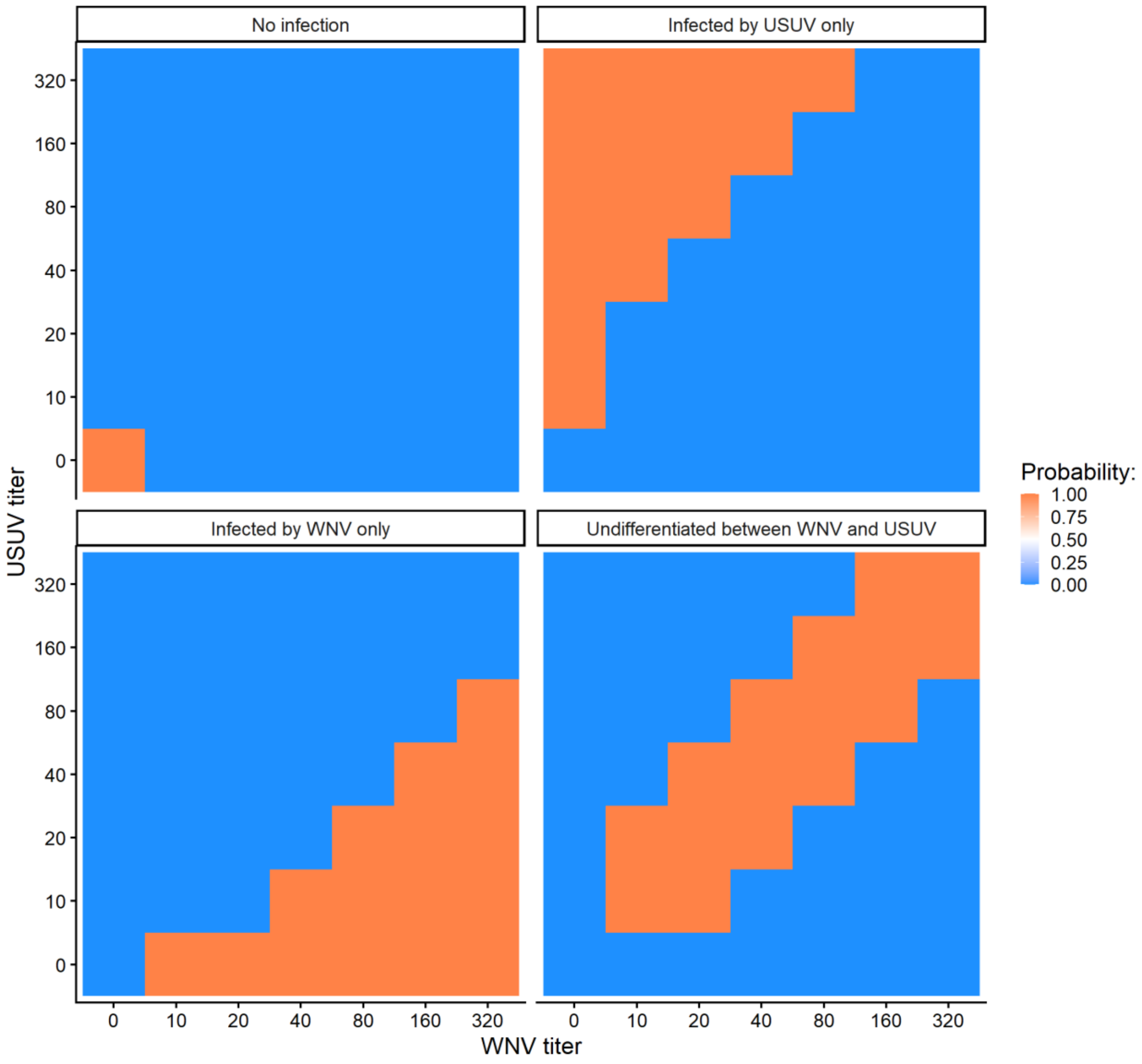


**Supplementary Figure S1.** Current threshold method used in the lab for classifying a serum based on VNT titer results for WNV and USUV (Chevalier et al. 2025).

**Supplementary Note 1. Details on the model components.**

Model parameters are summarized in Table 1 of the main manuscript.

1) <u>Epidemiological model</u>

The epidemiological model allowed to infer $I_{i,j,k}$, the true (unobserved) serological status for virus $k$ (coded 0 for negative and 1 for positive) at a sampling $j$ in an individual $i$. For any $i$, $k$ and $j \geq 1$, $I_{i,j,k}$ depended on a probability $p^{inf}_{i,j,k}$:

$$I_{i,j,k} \sim Bernoulli(p^{inf}_{i,j,k})$$

$$p^{inf}_{i,j,k} = \begin{cases} 1 - e^{-\lambda_{i,j,k}} \ if \ I_{i,j-1,k} = 0 \\ e^{-\tau(t_{i,j} - t_{i,j-1})} \ if \ I_{i,j-1,k} = 1 \end{cases}$$

Where $\tau$ was the seroreversion rate, assumed to be identical for WNV and USUV. We assumed that $I_{i,0,k}$, the true serological status on arrival at the zoo (birth or arrival from another zoo), was 0, meaning that we ignored maternal antibodies in newborn animals and viral exposure in other locations. The exception was a polar bear (*Ursus maritimus*) that was sampled on its arrival day at the zoo, was Panflavivirus ELISA-positive, and had a null WNV VNT titer and an USUV VNT titer of 20. It was considered truly positive to USUV on arrival at the zoo.

$\lambda_{i,j,k}$ was the cumulated force of infection of virus $k$ exerted on individual $i$ between $t_{i,j-1}$ and $t_{i,j}$, the times of samples $j$-$1$ and $j$ in individual $i$:

$$\lambda_{i,j,k} = \int_{t_{i,j-1}}^{t_{i,j}} \Lambda_k(t).\beta_i^{spec}.dt$$

Where $\beta^{spec}_i$ corresponded to the effect of the species category, with $\beta^{spec}_i=1$ if individual $i$ was a bird without water exposure, and $\beta^{spec}_i=\beta^{ma}$, $\beta^{spec}_i=\beta^{aqua}$, $\beta^{spec}_i=\beta^{stag}$ and $\beta^{spec}_i=\beta^{both}$ if it belonged to the category "Mammal or reptile", "Bird with aquatic lifestyle only", "Bird with proximity to stagnant water in summer nights only" and "Bird with both water exposures" respectively. $\Lambda_k(t)$ was the force of infection of virus $k$ on day $t$ that depended on the year $y(t)$:

$$\Lambda_k(t) = \begin{cases} \Lambda_{k,y(t)} \ if \ t \in [May\ 1^{st}; November\ 30^{th}] \\ 0 \ otherwise \end{cases}$$

2) <u>ELISA observation model</u>

For any sample $j$ in an individual $i$, the observed result of the Panflavivirus ELISA test, $F_{i,j}$, was modelled as:

$$F_{i,j} = \max(F_{i,j,WNV}, F_{i,j,USU})$$

Where, for each virus $k$:

$$F_{i,j,k} \sim Bernoulli(p^{panf}_{i,j,k})$$

$$p^{panf}_{i,j,k} = \begin{cases} \mu_{pan} \ if \ I_{i,j,k} = 1 \\ 1 - \gamma \ if \ I_{i,j,k} = 0 \end{cases}$$

Where $\mu_{pan}$ and $\gamma$ were respectively the sensitivity and specificity of the Panflavivirus ELISA test, assumed identical for both viruses.

3) VNT observation model

For any sample $j$ in any individual $i$, the (log2-transformed) titer resulting from the VNT for virus $k$, $V_{i,j,k}$, was modelled to first depend on a variable $D_{i,j,k}$ representing the detection (coded as 1) or not (coded as 0) of neutralizing antibodies directed against virus $k$. It was possible in case of positive true serological status for any of the two viruses:

$$D_{i,j,k} \sim Bernoulli(p_{i,j}^{VNT})$$

$$p_{i,j}^{VNT} = \begin{cases} \mu_{VNT}\ if\ I_{i,j,WNV} = 1\ or\ I_{i,j,USU} = 1 \\ 0\ if\ I_{i,j,WNV} = 0\ and\ I_{i,j,USU} = 0 \end{cases}$$

Where $\mu_{VNT} = \varepsilon.\mu_{pan}$ was the sensitivity of the VNT to the presence of antibodies against any of the two viruses, and VNT specificity was assumed to be 1. The distribution of $V_{i,j,k}$ was then a right-censored generalized Poisson distribution, as defined by (Harris et al. 2012), of mean $\eta_{i,j,k}$ and variance $\sigma^2$:

$$V_{i,j,k} \sim RC\_GenPoiss(\eta_{i,j,k}, \sigma^2)$$

Where $\sigma^2$ was set to 1.5 and where $\eta_{i,j,k}$ depended on (i) the titer value $\alpha_k$ associated to the presence of antibodies against virus $k$, (ii) the titer value associated to the presence of antibodies against the other virus, due to the serological cross-reactivity, and (iii) a parameter $\psi$ quantifying the strength of the serological cross-reactivity between the two viruses:

$$\eta_{i,j,WNV} = \delta + D_{i,j,WNV}.(\alpha_{WNV}.I_{i,j,WNV} + \psi_{USU-WNV}.\alpha_{USU}.I_{i,j,USU})$$

$$\eta_{i,j,USU} = \delta + D_{i,j,USU}.(\alpha_{USU}.I_{i,j,USU} + \psi_{WNV-USU}.\alpha_{WNV}.I_{i,j,WNV})$$

Where including $\delta = 0.001$ was in order to ensure $\eta_{i,j,k} > 0$.

This distribution was right-censored because VNT titer values above 320 (or 6 when log2-transformed) were not observed.

**Supplementary Note 2. Derivation of prior distributions for the ELISA sensitivity $\mu_{pan}$ and specificity $\gamma$, and for the ratio of sensitivity between VNT and ELISA $\varepsilon$, based on the literature.**

We used informative priors for some of the model parameters, based on the literature.

First, for the seroreversion rate parameter ($\tau$), we used a fixed value that was estimated by combining a model to serological data collected in Guadeloupe (Caribbean), as part of a previous study in our group (Hamouche et al. 2025).

Second, we estimated distributions for the ELISA sensitivity and specificity, and ratio of sensitivity of the VNT as compared to the ELISA test ($\varepsilon$), based on (Girl et al. 2024). In this study, the authors tested the same set of samples with different serological tests, including a WNV-ELISA test and a WNV-NT (virus neutralization test). We used their results to fit the following model:

$$Res_i^{ELISA} \sim Bernoulli(p_i^{ELISA})$$

$$Res_i^{NT} \sim Bernoulli(p_i^{NT})$$

$$p_i^{ELISA} = Pos_i . Sens_{ELISA} + (1 - Pos_i).(1 - Spec_{ELISA})$$

$$p_i^{NT} = Pos_i . \varepsilon . Sens_{ELISA} + (1 - Pos_i).(1 - Spec_{NT})$$

$$Pos_i \sim Bernoulli(Prev)$$

Where $Res^{NT}_i$ and $Res^{ELISA}_i$ were the results (coded 1 for positive and 0 for negative) of the NT and ELISA test in sample i respectively, observed in the study. $Sens_{ELISA}$ and $Spec_{ELISA}$ were the sensitivity and specificity of the ELISA test, $Spec_{NT}$ was the specificity of the NT, assumed equal to 0.999 (Hamouche et al. 2025), and $\varepsilon$ was as defined in the main manuscript. $Pos_i$ was the true (unobserved) seropositivity to WNV in sample i. Therefore, we estimated distributions of $Sens_{ELISA}$, $Spec_{ELISA}$ and $\varepsilon$, with (Girl et al. 2024) data, and subsequent median estimates and 95% credible intervals. We fitted a Beta distribution to the distribution of $\varepsilon$ and used it as a prior distribution in the current study.

Third, another study (Hogrefe et al. 2004) tested serums with a WNV IgG ELISA test, including blood donor samples collected from areas that had not been affected by WNV yet, representing true negatives to assess the test's specificity. Out of 236 samples, 7 resulted in a positive result, 4 in an equivocal result and 225 in a negative result. When considering equivocal results as positives, we could then estimate the specificity of the WNV IgG ELISA test to 95.3% (95% confidence interval: [91.6% ; 97.5%], Clopper-Pearson method).

A fourth study (Beck et al. 2017) had the same set of samples tested by multiple laboratories, with various serological testing methods, including WNV IgG ELISA and VNT methods. From this publication, we used the test results to the true WNV-negative samples (samples S1, S2, S3 and S4 in 24 laboratories in 2010, and sample S1 in 31 laboratories in 2013) to compute a global specificity for the IgG ELISA test (99.2% [95.0% ; 99.9%]). Similarly, we used results to the true WNV-positive samples (samples S13 and S16 in 24 laboratories in 2010, and samples S5, S6 and S7 in 31 laboratories in 2013) to compute the IgG ELISA test sensitivity (96.5% [91.5% ; 98.7%]).

Finally, for ELISA sensitivity and specificity, we combined these previously estimated point estimates and intervals to fit Beta distributions and used them as informative prior distributions in the current study (Supplementary Figure S2).

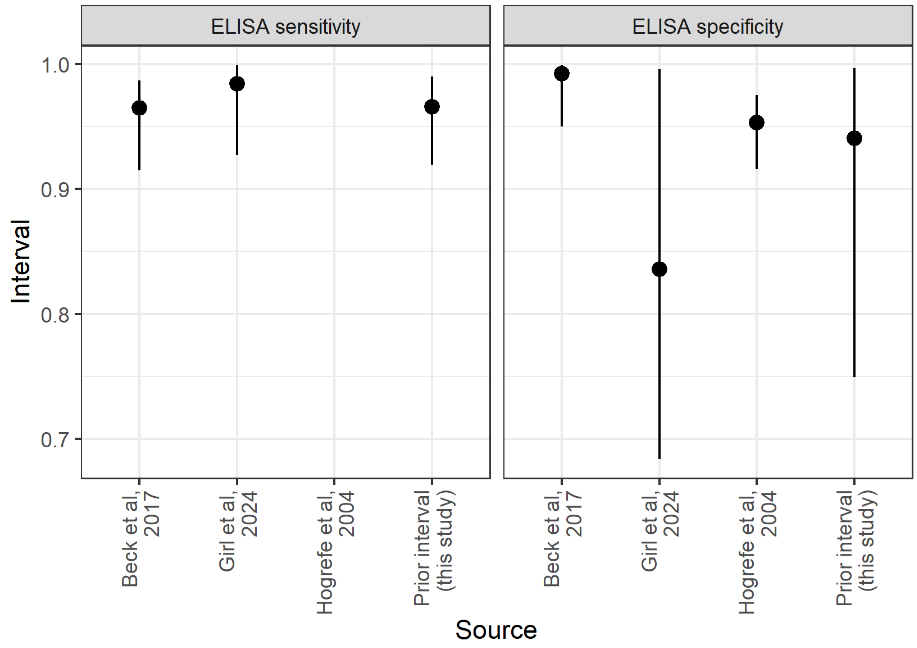


**Supplementary Figure S2.** Prior distributions used for the ELISA test sensitivity ($\mu_{pan}$) and specificity ($\gamma$) in the current study, computed from point estimates and intervals derived from previous studies (see details in Supplementary Note 2).

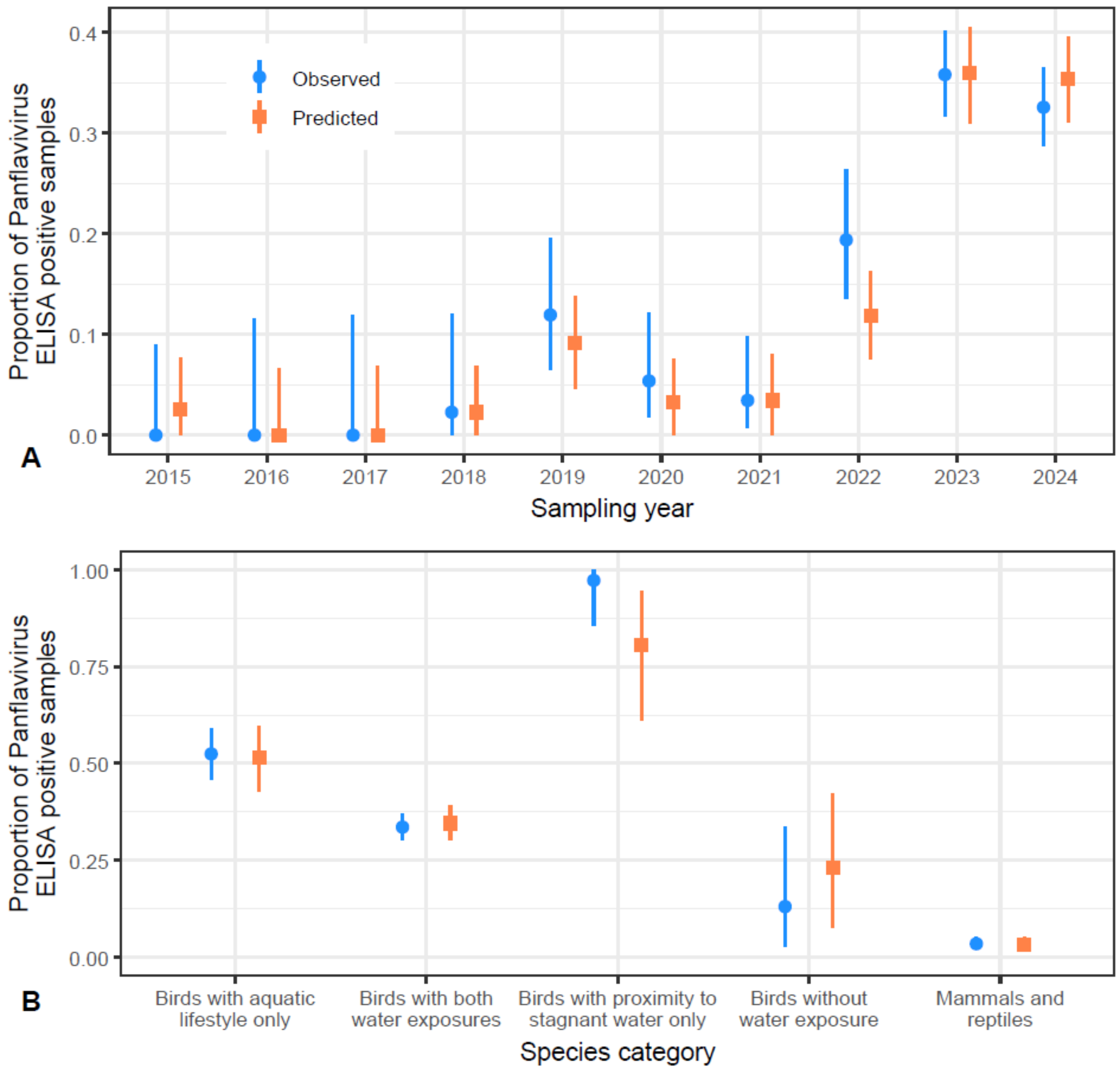


**Supplementary Figure S3.** Goodness of fit of the model to Panflavivirus ELISA data. Proportion of positive Panflavivirus ELISA results by year of sampling (panel A) and by species category (panel B), as observed in sampled zoo animals (blue circles) and as predicted by the fitted model (orange squares). Intervals are 95% confidence interval of the proportion (Clopper-Pearson method) for observed results, and 95% prediction intervals across 1000 repetitions of the model for predictions.

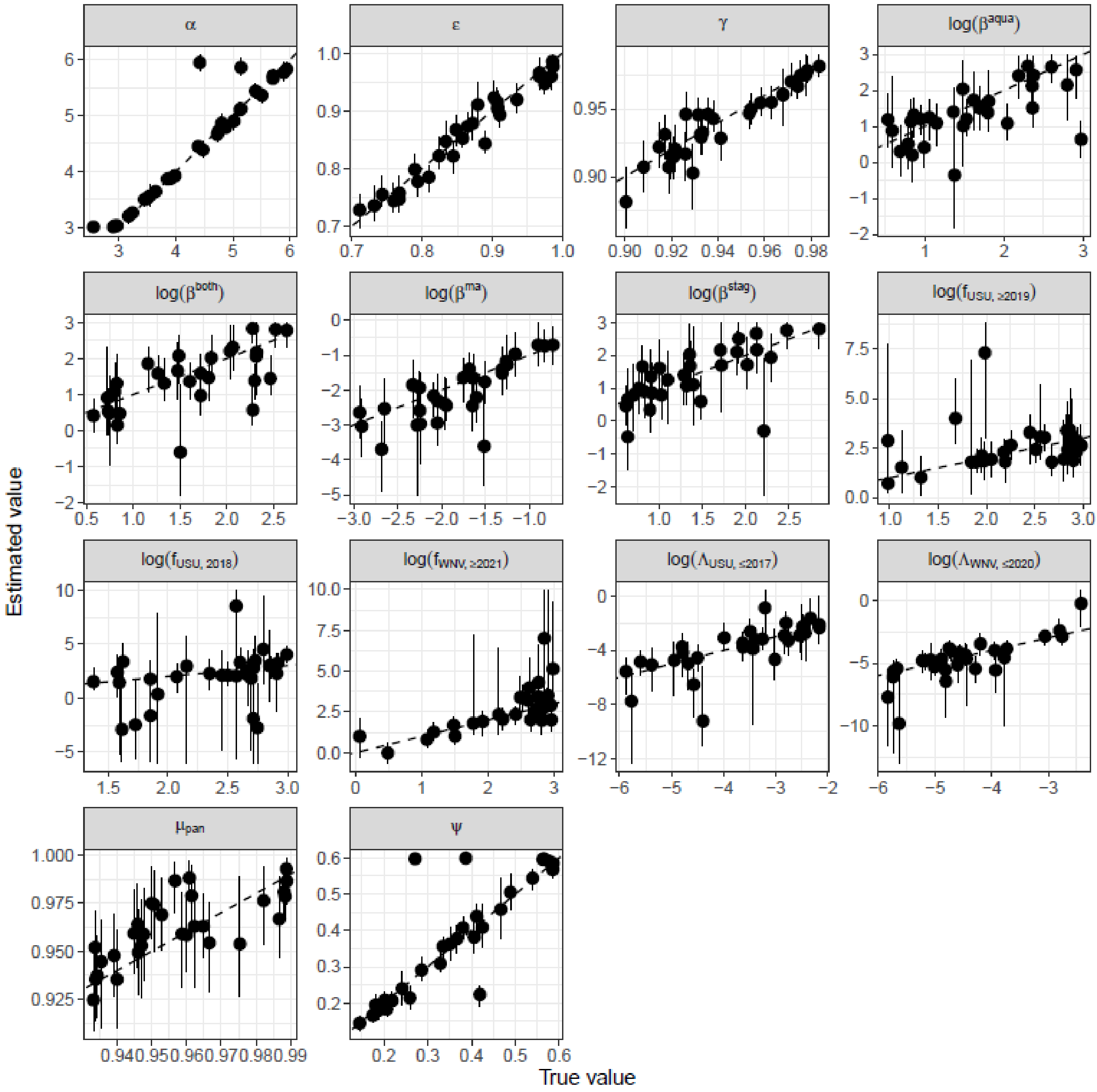


**Supplementary Figure S4.** Results of the model validation using 30 synthetic datasets. Mock (synthetic) data was first generated using known ("true") values for the parameters, and the model was then fitted to the synthetic data. Points and intervals correspond to the posterior median and 95% credible interval of the parameters.

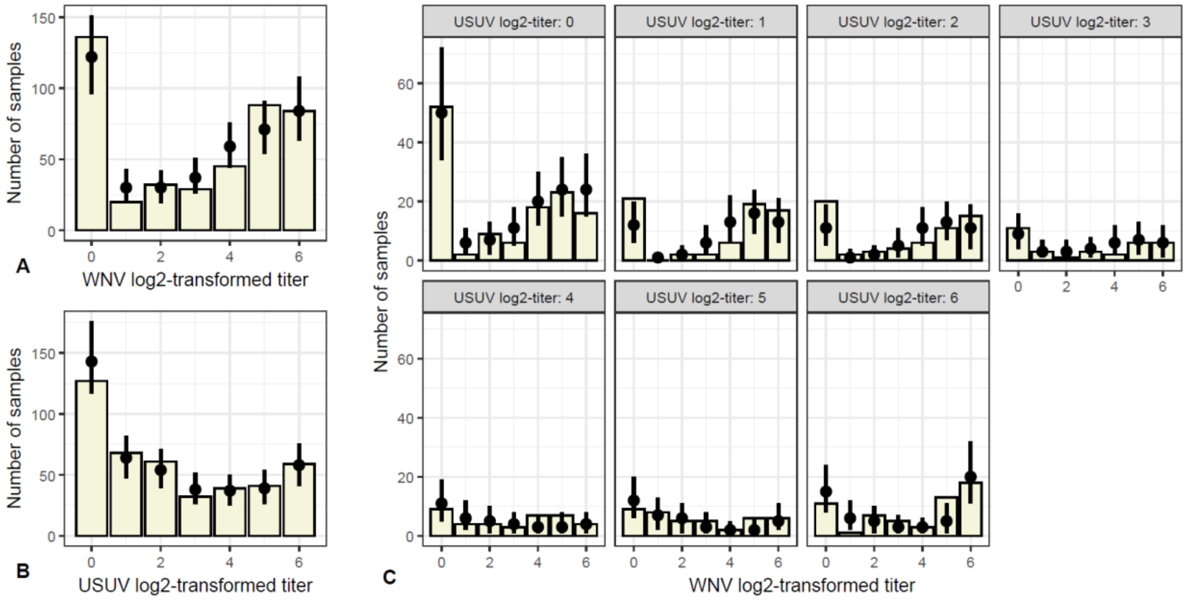


**Supplementary Figure S5.** Goodness of fit of the model to Virus Neutralization Test data. Marginal (panels A and B) and joint (panel C) distributions of the observed (bars) vs. predicted (point and interval, representing the median and 95% prediction interval of 1000 model repetitions) titer values for WNV and USUV.

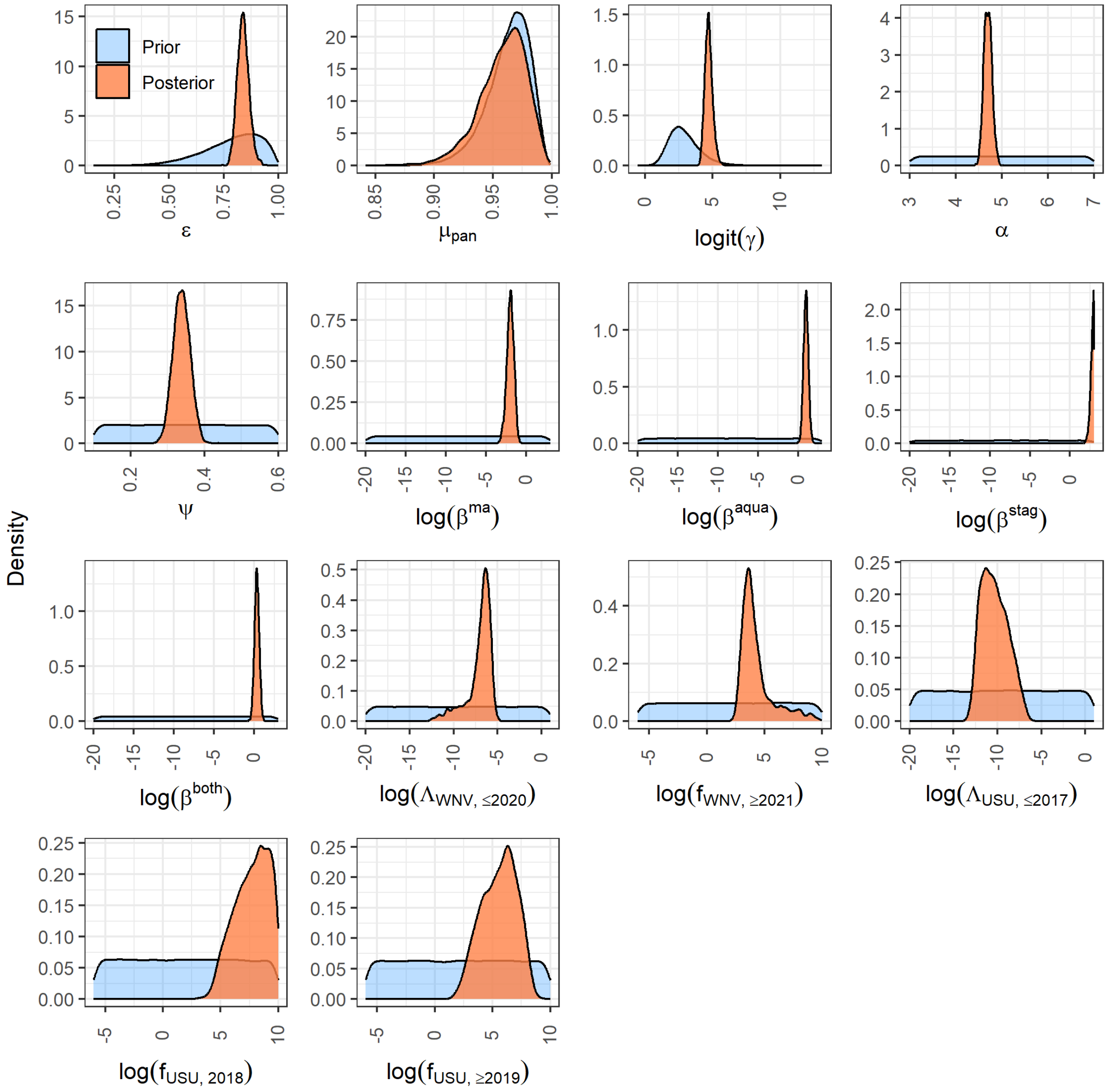


**Supplementary Figure S6.** Prior and posterior distributions of the model parameters.

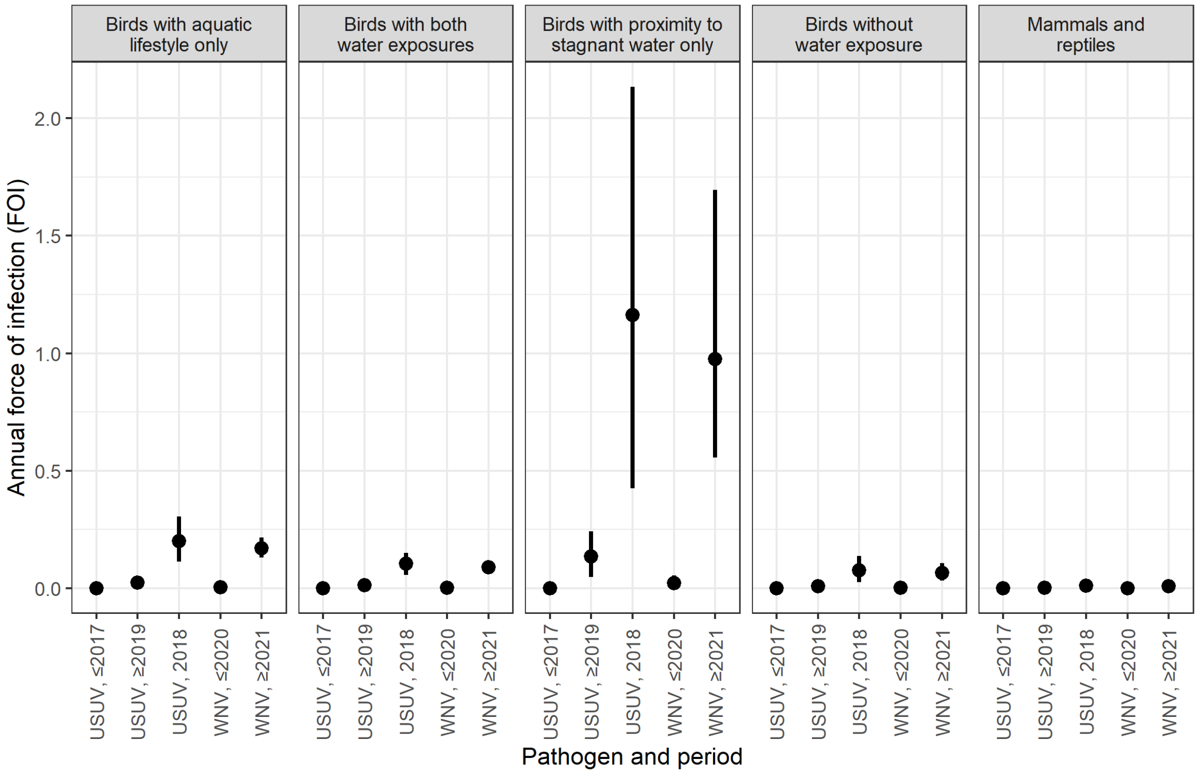


**Supplementary Figure S7.** Posterior estimates (median and 95% credible interval) of the annual force of infection (FOI) by USUV and WNV at the different periods in the zoo, in the different species categories. We considered the exposure time was only between May 1$^{st}$ and November 30$^{th}$ of each year (the mosquito season).

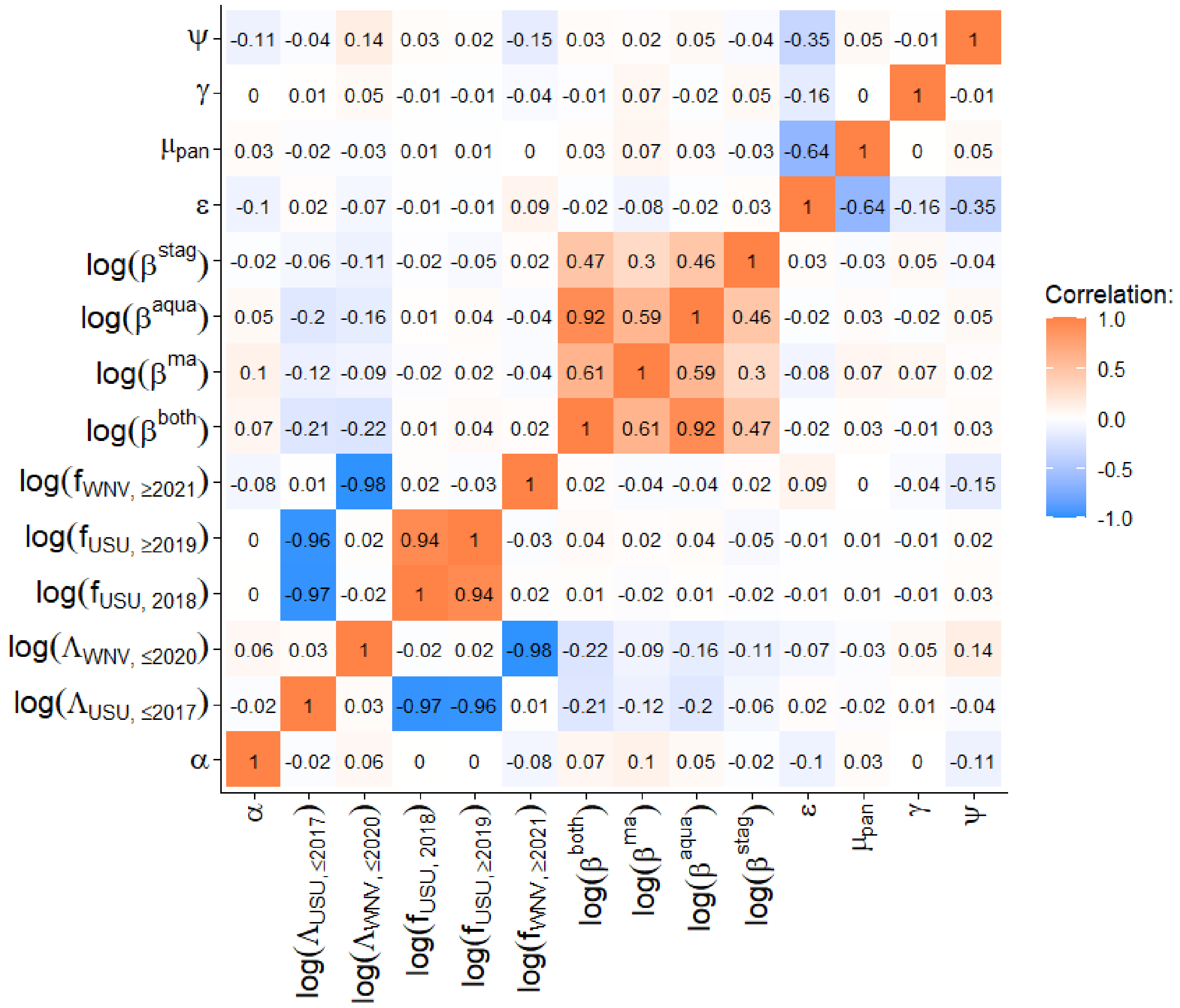


**Supplementary Figure S8.** Correlation between posterior distributions of model parameters.

**Supplementary Note 3. Model-based diagnosis of flavivirus seropositivity.**

Using our model parameter estimates, we computed the probability for a sample s to have a positive true serostatus for WNV (i.e. $I_{s,WNV} = 1$) and USUV (i.e. $I_{s,USU} = 1$), given values of the titer for both viruses ($V_{s,WNV}$ and $V_{s,USU}$). Taking notations from the Supplementary Note 1, we could calculate for $n_{WNV}$, $n_{USU}$ $\in \{0; 1; 2; 3; 4; 5; 6\}$ and a, b $\in \{0; 1\}$:

$$P\left(I_{s,WNV} = a, I_{s,USU} = b \middle| V_{s,WNV} = n_{WNV}, V_{s,USU} = n_{USU}\right)$$

$$= \frac{P\left(V_{s,WNV} = n_{WNV}, V_{s,USU} = n_{USU} \middle| I_{s,WNV} = a, I_{s,USU} = b\right). P\left(I_{s,WNV} = a, I_{s,USU} = b\right)}{P\left(V_{s,WNV} = n_{WNV}, V_{s,USU} = n_{USU}\right)}$$

$$= \frac{P\left(V_{s,WNV} = n_{WNV}, V_{s,USU} = n_{USU} \middle| I_{s,WNV} = a, I_{s,USU} = b\right). P\left(I_{s,WNV} = a, I_{s,USU} = b\right)}{\sum_{x \in \{0;1\}} \sum_{y \in \{0;1\}} P\left(I_{s,WNV} = x, I_{s,USU} = y\right). P\left(V_{s,WNV} = n_{WNV}, V_{s,USU} = n_{USU} \middle| I_{s,WNV} = x, I_{s,USU} = y\right)}$$

In practice, we drawn 1000 samples from the joint posterior distribution to get values of the parameters and proportions of positive true serostatus in the whole dataset (for WNV only, USUV only, both viruses and none of the viruses), and derived the median probabilities.

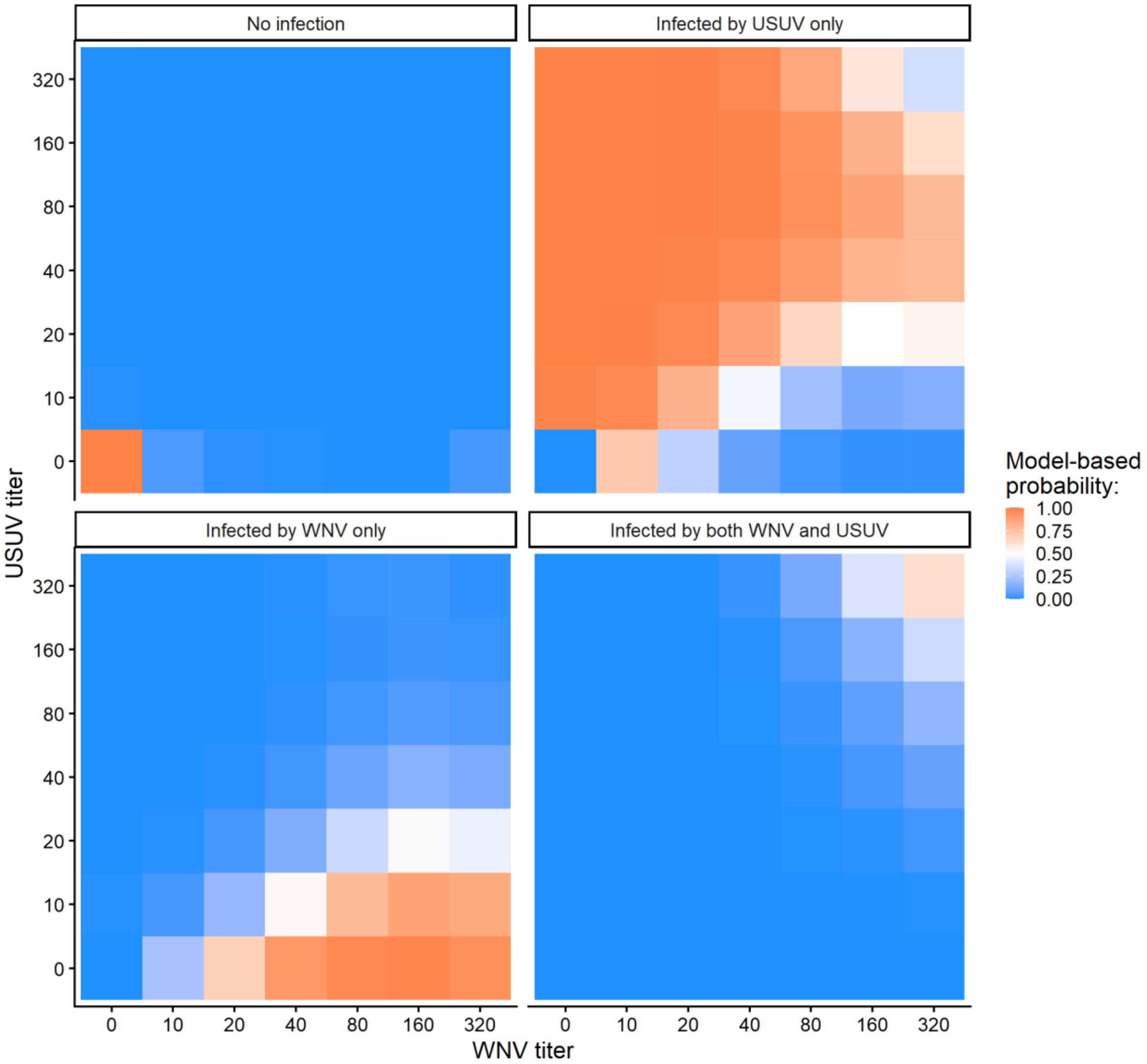


**Supplementary Figure S9.** Model-based probability of true serological status (past infection) of a sample to USUV and/or WNN, given VNT titer values for both viruses and assuming a global true seroprevalence (i.e. the proportion of positive true serostatus) of 15% for USUV and 0.5% for WNV (see details on the computation in Supplementary Note 3).

**Supplementary Table S4.** Deviance Information Criterion (DIC) for the main model and for alternative models tested as part of the sensitivity analysis.

| **Model assumptions** | **DIC value** |
|---|---|
| $\alpha_{WNV} = \alpha_{USU}$, $\psi_{WNV\text{-}USU} = \psi_{USU\text{-}WNV}$, three circulation periods for USUV and two for WNV (main analysis) | 1,694,404 |
| $\alpha_{WNV} \neq \alpha_{USU}$, $\psi_{WNV\text{-}USU} = \psi_{USU\text{-}WNV}$, three circulation periods for USUV and two for WNV | 1,694,426 |
| $\alpha_{WNV} = \alpha_{USU}$, $\psi_{WNV\text{-}USU} \neq \psi_{USU\text{-}WNV}$, three circulation periods for USUV and two for WNV | 1,694,428 |
| $\alpha_{WNV} = \alpha_{USU}$, $\psi_{WNV\text{-}USU} = \psi_{USU\text{-}WNV}$, four circulation periods for USUV and two for WNV | 1,694,396 |
| $\alpha_{WNV} = \alpha_{USU}$, $\psi_{WNV\text{-}USU} = \psi_{USU\text{-}WNV}$, three circulation periods for USUV and one for WNV | 1,694,426 |
| $\alpha_{WNV} = \alpha_{USU}$, $\psi_{WNV\text{-}USU} = \psi_{USU\text{-}WNV}$, three circulation periods for USUV and three for WNV | 1,694,409 |
| $\alpha_{WNV} = \alpha_{USU}$, $\psi_{WNV\text{-}USU} = \psi_{USU\text{-}WNV}$, three circulation periods for USUV and two different periods for WNV (pre-2021 and post-2022) | 1,694,415 |
| $\alpha_{WNV} = \alpha_{USU}$, $\psi_{WNV\text{-}USU} = \psi_{USU\text{-}WNV}$, three circulation periods for USUV and two for WNV, different values of $\beta^{aqua}$, $\beta^{stag}$ and $\beta^{both}$ for USUV *vs.* WNV | 1,694,416 |

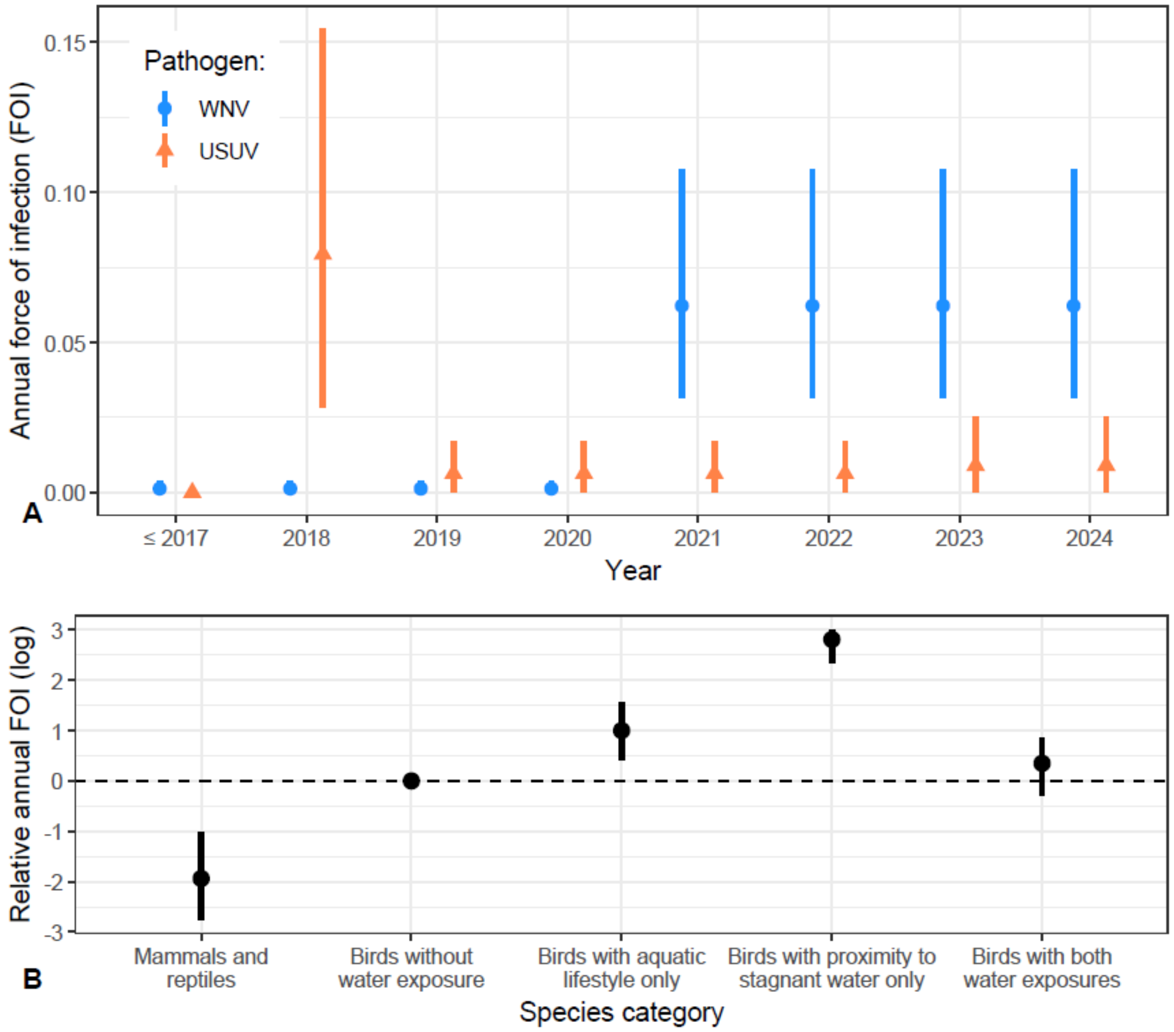


**Supplementary Figure S10.** Model results in an alternative model that assumes four, instead of three, circulation periods for USUV. Panel A: Posterior estimates (median and 95% credible interval) of $\Lambda_{k,y(t)}$, the annual forces of infection (FOI) in birds with no water exposure, for WNV and USUV, in La Palmyre zoo. Panel B: Posterior estimates (median and 95% credible interval) of the log-scaled relative risk of infection in mammals and reptiles, and in birds with exposure to water (aquatic lifestyle and/or proximity to stagnant water in summer nights) as compared to birds with no water exposure (respectively $\beta^{ma}$, $\beta^{aqua}$, $\beta^{stag}$ and $\beta^{both}$).

### Bibliography

Beck, Cécile, Steeve Lowenski, Benoit Durand, Céline Bahuon, Stéphan Zientara, and Sylvie Lecollinet. 2017. "Improved Reliability of Serological Tools for the Diagnosis of West Nile Fever in Horses within Europe." *PLOS Neglected Tropical Diseases* 11 (9): e0005936. https://doi.org/10.1371/journal.pntd.0005936.

Chevalier, Noémie, Camille V. Migné, Teheipuaura Mariteragi-Helle, et al. 2025. "Seroprevalence of West Nile, Usutu and Tick-Borne Encephalitis Viruses in Equids from South-Western France in 2023." *Veterinary Research* 56 (1): 1. https://doi.org/10.1186/s13567-025-01508-w.

Girl, Philipp, Kathrin Euringer, Mircea Coroian, Andrei Daniel Mihalca, Johannes P. Borde, and Gerhard Dobler. 2024. "Comparison of Five Serological Methods for the Detection of West Nile Virus Antibodies." *Viruses* 16 (5): 5. https://doi.org/10.3390/v16050788.

Hamouche, Celia, Jennifer Pradel, Nonito Pagès, et al. 2025. "Reconstructing the Silent Circulation of West Nile Virus in a Caribbean Island during 15 Years Using Sentinel Serological Data." *PLOS Neglected Tropical Diseases* 19 (6): e0012895. https://doi.org/10.1371/journal.pntd.0012895.

Harris, Tammy, Zhao Yang, and James W. Hardin. 2012. "Modeling Underdispersed Count Data with Generalized Poisson Regression." *The Stata Journal* 12 (4): 736–47. https://doi.org/10.1177/1536867X1201200412.

Hogrefe, Wayne R., Ronald Moore, Mary Lape-Nixon, Michael Wagner, and Harry E. Prince. 2004. "Performance of Immunoglobulin G (IgG) and IgM Enzyme-Linked Immunosorbent Assays Using a West Nile Virus Recombinant Antigen (preM/E) for Detection of West Nile Virus- and Other Flavivirus-Specific Antibodies." *Journal of Clinical Microbiology* 42 (10): 4641–48. https://doi.org/10.1128/JCM.42.10.4641-4648.2004.
